# An explanation for SARS-CoV-2 rebound after Paxlovid treatment

**DOI:** 10.1101/2023.05.30.23290747

**Authors:** Alan S. Perelson, Ruy M. Ribeiro, Tin Phan

## Abstract

In a fraction of SARS-CoV-2 infected individuals treated with the oral antiviral Paxlovid, the virus rebounds following treatment. The mechanism driving rebound is not understood. Here, we show that viral dynamic models based on the hypothesis that Paxlovid treatment near the time of symptom onset halts the depletion of target cells, but may not fully eliminate the virus, which can lead to viral rebound. We also show that the occurrence of viral rebound is sensitive to model parameters, and the time treatment is initiated, which may explain why only a fraction of individuals develop viral rebound. Finally, the models are used to test the therapeutic effects of two alternative treatment schemes. These findings also provide a possible explanation for rebounds following other antiviral treatments for SARS-CoV-2.

**Significance:** Paxlovid is an effective treatment for SARS-CoV-2. In some patients treated with Paxlovid, the initial reduction in viral load is followed by a rebound once treatment is stopped. Understanding the mechanisms of the rebound may help us develop better treatment strategies to minimize this possibility. We hypothesize that early treatment with Paxlovid stops viral growth, but may not fully clear the virus, thus preserving host resources that would have otherwise been used by the virus. Once treatment ends, the remaining viruses can utilize the available resources to grow, leading to the observed transient viral rebound. We built standard viral dynamic models based on this hypothesis and fit the models to data to show its feasibility. We further examined the effect of two alternative treatment schemes.

## Introduction

A 5-day course of Paxlovid is recommended for individuals who tested positive for SARS-CoV-2 with mild to moderate symptoms and a high risk of progression to severe disease (1, 2). Treatment is suggested to be initiated as soon as possible and within 5 days of symptom onset, with two doses per day. A dose of Paxlovid consists of 300 mg nirmatrelvir and 100 mg of ritonavir, where nirmatrelvir is a protease inhibitor that blocks SARS-CoV-2 replication and ritonavir reduces the liver catabolism of nirmatrelvir and thus prolongs its half-life (1). While Paxlovid substantially reduces the risk of progression to severe COVID-19 and can shorten the duration of infectiousness in high-risk individuals (2-5), in some cases viral rebound and recurring symptoms occur after the 5-day treatment, including in individuals who have been vaccinated and/or boosted. Symptoms resolve within a median of 3 days after rebound without additional intervention (6). Some individuals with viral rebound are reported to have culturable virus up to 16 days after the initial diagnosis (7) and it is possible that transmission to close contacts may occur during the rebound period (8). Furthermore, viral rebound does not appear to be caused by the emergence of drug resistant mutants (8-10).

The fraction of individuals treated with Paxlovid that exhibit viral rebound has not been well quantified, and estimates vary. For example, in a cohort of 484 high-risk individuals treated with Paxlovid, four individuals (0.8%) experienced rebound of symptoms (11). In the phase 3 Paxlovid clinical trial, EPIC-HR, the fraction of individuals with viral rebound (positive PCR test) and recurring symptom was 1-2% (2). A recent study at the Beth Israel Deaconess Medical Center reported 3 out of 11 mRNA vaccinated individuals who were treated with Paxlovid developed rebound (27% vs. 4% in an untreated group) (12). In a retrospective study of electronic health records of 11,270 patients aged 18 years or older that contracted COVID-19 between 1/1/2022 and 6/8/2022 and were treated with Paxlovid, 3.53% exhibited rebound between 2 and 7 days after treatment (13).

In this study, we analyze the data presented in Charness et al. (8), where quantitative PCR data is available for three individuals who experienced viral and symptom rebounds after taking Paxlovid. In all three individuals, no mutations occurred during treatment in the gene encoding the protease targeted by nirmatrelvir and there was no evidence of reinfection by a different variant (8). This is like other studies reporting no detectable mutations in Paxlovid-treated rebound individuals (7, 10). We show that viral dynamic models explain the rebound phenomenon, based on the idea that 5-day Paxlovid treatment started near the time of symptom onset reduces the depletion of target cells but does not fully eliminate virus, thus allowing the virus to rebound once treatment is stopped. Further, we show that such models generate viral dynamic profiles that agree with the data. We also show that the occurrence of viral rebound is sensitive to model parameters and the time therapy is started so that one would expect only a minority of treated individuals to rebound. Lastly, we use our model to examine the potential effects of giving Paxlovid for 10 days or a second course of Paxlovid after symptoms return. While we use Paxlovid as a case study, our theory also can also explain the viral rebound observed after treatment with molnupiravir (13), the only other oral antiviral with emergency use authorization.

## Results

### The viral dynamic model matches the viral rebound observed in patient data

Our viral dynamic model (see Methods) can describe the observed data including the viral rebound in the three treated individuals (Fig. 1). We observe that after treatment starts, the decline in target cells is temporarily halted (Fig. 1), which indicates Paxlovid preserves target cells. The remaining target cells can support viral replication if there is viable virus remaining after treatment ends. Note that in patient 1 the virus rebounds to a very high level which depletes the remaining target cells. Due to this there was no support for including an adaptive immune response in the model for this patient. In the other two patients, ultimate clearance of the virus was due to an adaptive immune response developing at time *t*\* specified in the figure caption.

**Fig. 1.**
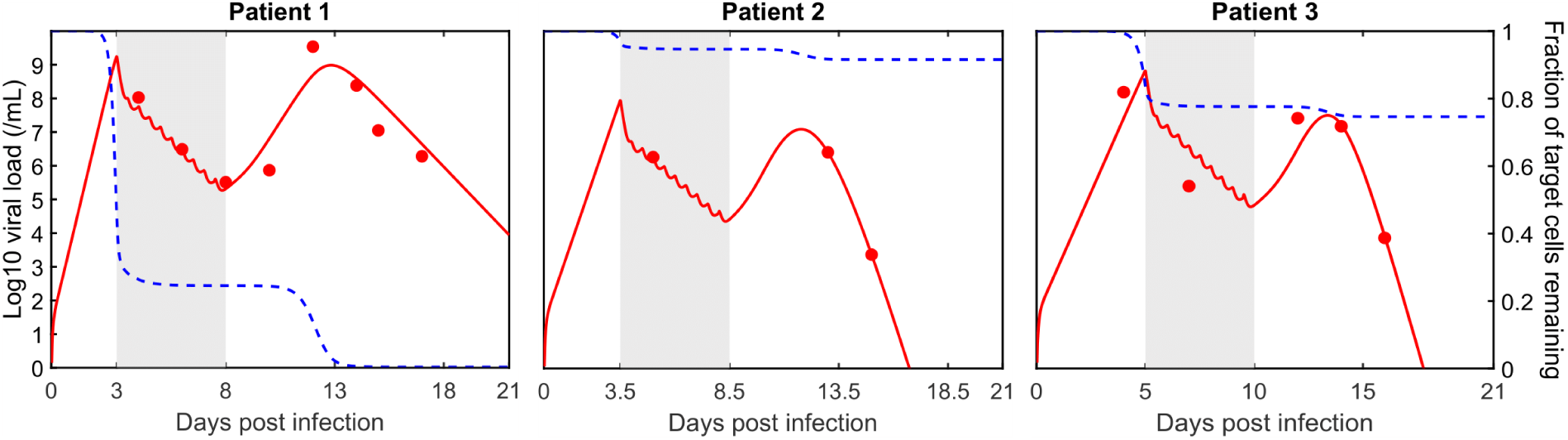
Best fit of the viral dynamic model (solid curves) to the viral load data (open circles). The shaded area is the duration of treatment. The dashed curve is the model predicted fraction of target cells remaining. The following parameter were fixed for all three patients at values previously determined in the literature (14): *k* = 4 day^−1^, *δ* = 1.7 day^−1^, *c* = 10 day^−1^. Additionally, fitting indicated for patient 1, *β* = 2.32 × 10^−9^ mL RNA copies^−1^day^−1^, *π* = 1.77 × 10^3^ RNA *c*opies mL^−1^day^−1^, and treatment was from day 3 to day 8. For patient 2, *β* = 1.54 × 10^−9^ mL RNA copies^−1^day^−1^, *π* = 1.47 × 10^3^ RNA copies mL^−1^day^−1^, *t*^∗^ = 9.1 days, *σ* = 0.82 day^−1^ and treatment was from day 3.5 to day 8.5. For patient 3, *β* = 8.24 × 10^−10^ mL RNA copies^−1^day^−1^, *π* = 1.89 × 10^3^ RNA copies mL^−1^day^−1^, *t*^∗^ = 12.0 days, *σ* = 1.22 day^−1^ and treatment was from day 5 to day 10.

### The effects of two hypothetical alternative Paxlovid treatment schedules

Although extending the treatment duration of Paxlovid is not currently recommended as the vast majority of treated individuals do not rebound, it is still of interest to see what our model predicts if we extend the treatment duration by either having one continuous 10-day treatment or start an additional 5-day treatment one day after symptoms rebounded. The times symptoms returned were 5, 5 and 3 days after treatment ended for patients 1, 2, and 3, respectively (8).

By extending the treatment duration, it is possible to prevent viral rebound in patients 2 and 3 (Fig. 2A). However, for patient 1, viral rebound is predicted to still occur after the end of treatment as the viral load and number of infected cells were not driven to extinction. For patients 2 and 3 adaptive immunity starting between days 9 and 12, as predicted in Fig.1, was needed to explain the observed data and in the model simulations drove the viral load to undetectable levels before the extended duration treatment ended. On the other hand, adding a second course of treatment one day after symptoms returned is predicted to have minimal impact on the viral trajectory toward clearance compared to a single course of treatment especially for patients 1 and 2 (Fig. 2B).

**Fig. 2.**
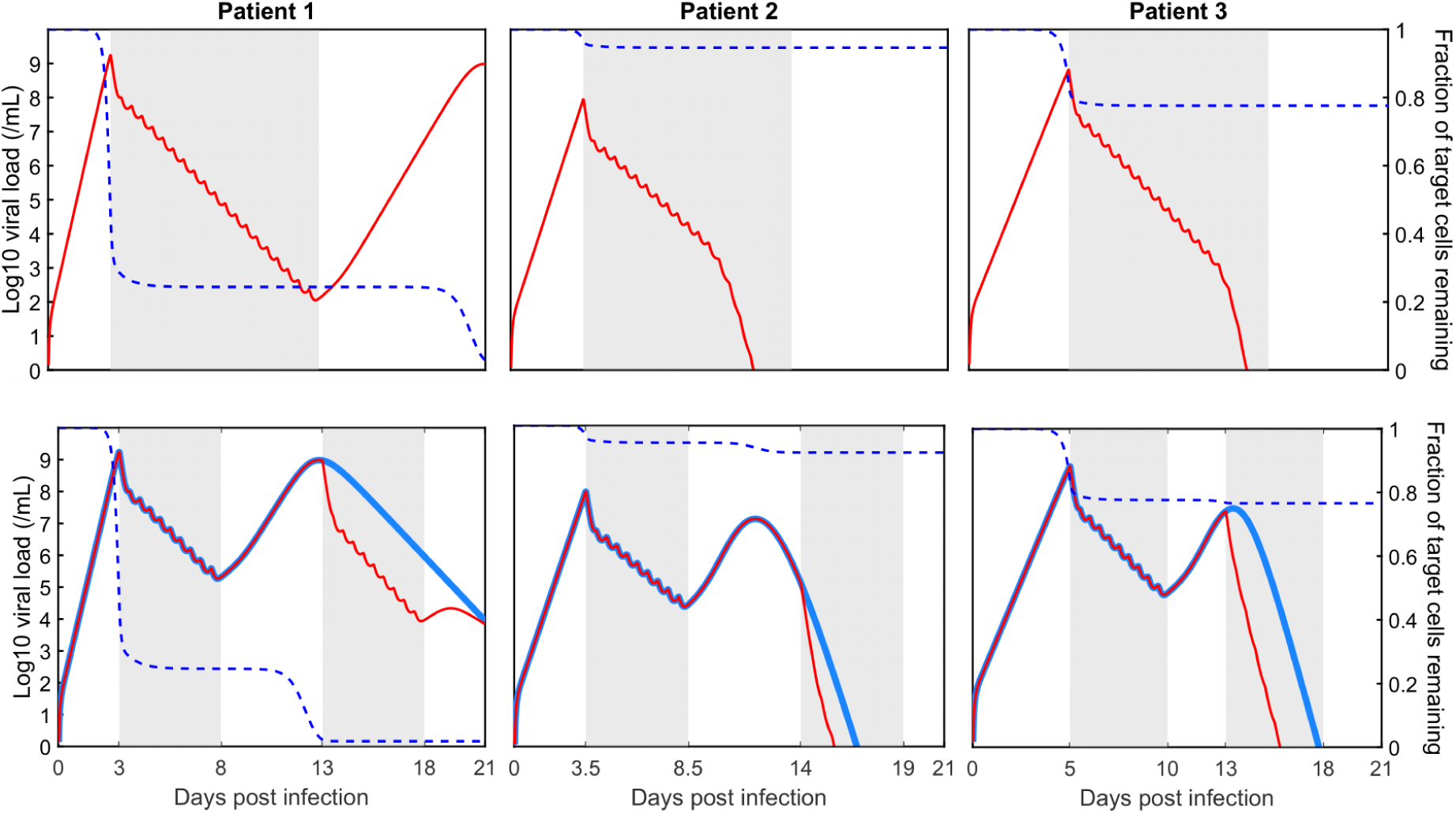
Model simulation with a 10-day hypothetical treatment: (A) 10 consecutive days treatment (B) a second Paxlovid course started one day after symptoms returned. Solid red curves are the model predicted viral load using the parameters given in the Fig. 1 caption. The dashed blue curve is the model predicted number of target cells under the hypothetical treatment. The solid blue curves in (B) are the predicted viral trajectory with the standard five-day course of Paxlovid reproduced from Fig. 1. The shaded area is the duration of treatment.

### The sensitivity of viral rebound to treatment initiation time and patient specific parameters

We used simulation experiments to show that delaying treatment with Paxlovid can decrease the probability of rebound in general. We first generated an *in silico* cohort of SARS-CoV-2 patients as described in the Supplementary text. We then simulated a trial with 100 *in silico* treated patients and assessed what percentage exhibited rebound, which was defined as the viral load returning above 10^5^RNA copies per mL The rationale for 10^5^RNA copies per mL is that above this value infectious cultures can often be obtained, indicating the possibility of transmission. We repeated this trial protocol 20 times each for treatment starting at days 2, 3, 4 and 5. In Fig. 3A, we present boxplots of the percentage of rebound patients obtained from these 20 trials. Examples of 100 viral trajectories for each treatment initiation time are provided in Fig. 3B. Based on these *in silico* trial results, the model predicts the probability of rebound decreases quickly with increasing delay in Paxlovid initiation. We also obtained a similar conclusion for the second model we developed that included an innate immune response (see Methods), see Fig. S9 in the *Supplementary Materials*.

**Fig. 3.**
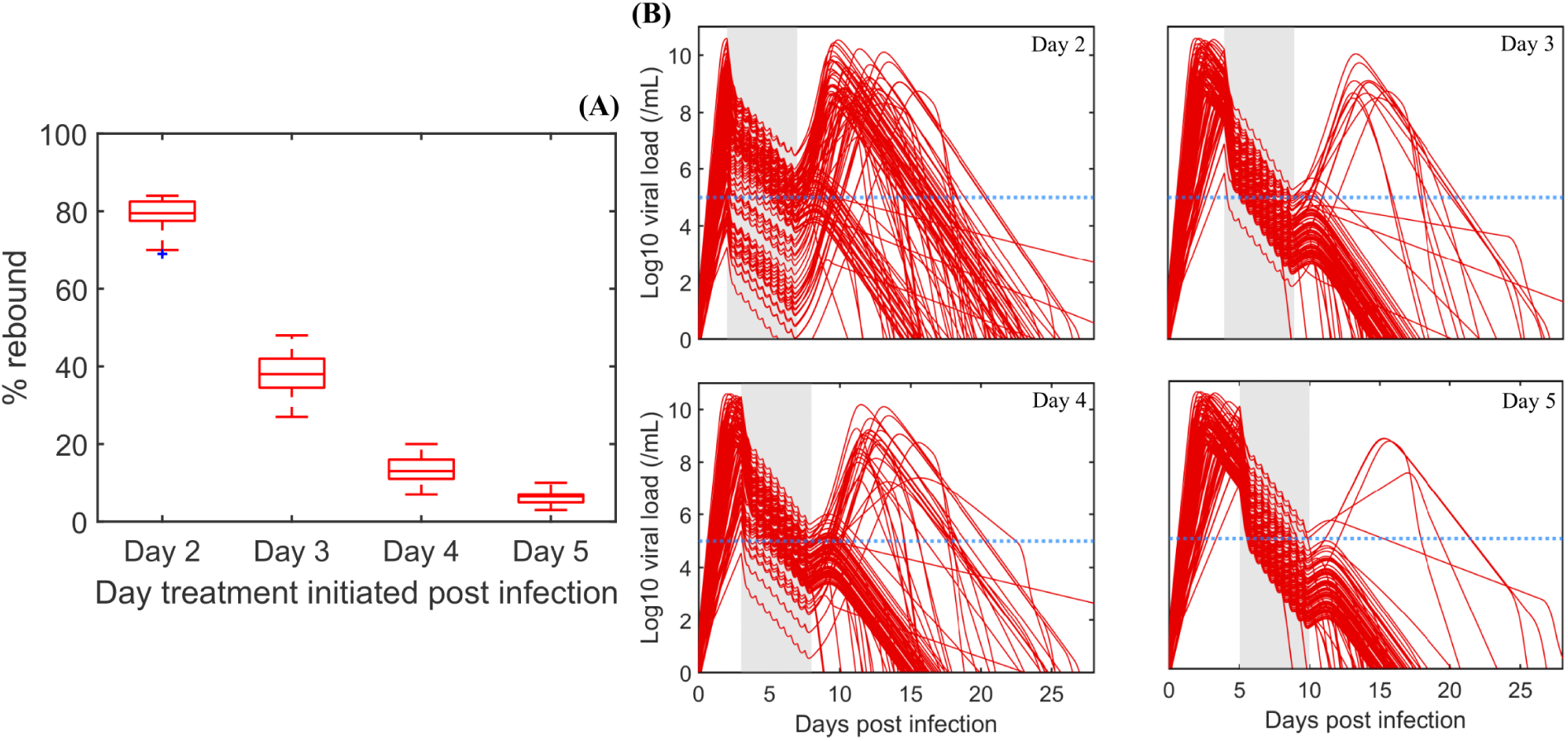
Percentage of predicted *in silico* patients with rebound relative to the time of treatment initiation. (A) Boxplots of the percentage of rebound cases from 20 trials with 100 virtual patients each for a different treatment initiation time. (B) Examples of 100 viral trajectories. Note the decreasing number of rebound cases as treatment starts later. The horizontal blue dashed line is the viral load threshold used to define rebound.

Another factor that may affect the level of target cells at the end of treatment are viral dynamic parameters: the infection rate *β*, the viral production rate *π*, and the viral clearance rate *c*. In Fig. 4, we simulate our viral dynamic model with parameters for patient 1, the patient with the most data, but varying values for *β*, *π* or *c*. When either the infection rate or the viral production rate is higher by 15%, virus grows faster, leading to a faster reduction in the available target cells and no rebound. The same occurs if the viral clearance rate is reduced by ∼20%. At the end of treatment, less than 1.8% of target cells remain with an increased value of either *β* or *π* by 15% or a decrease in *c* by 20% versus 24.4% remaining with the baseline parameters (Figs. 1 and 2). We further generalized the effect of varying the viral dynamics parameters on rebound using the *in silico* patients (Fig. S7, *Supplementary Materials*). Altogether, we showed that differences among treated individuals in viral dynamic parameters may explain why some individuals may be more likely to experience rebound than others.

**Fig. 4.**
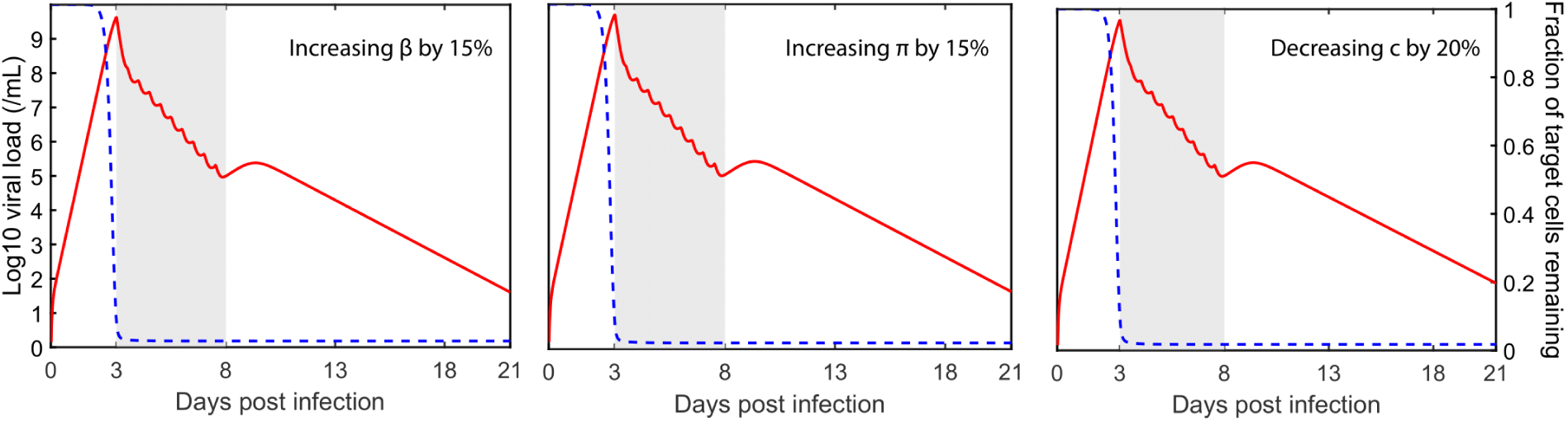
Model simulation with variation in values of *β*, *π* and *c* for patient 1. Solid red curves are the model simulated viral load. The dashed blue curve is the number of target cells. The shaded area is the duration of treatment. In the panels from left to right, the model is simulated with *β* 15% higher, *π* 15% higher, and c 20% lower than in the best-fit in Fig. 1 for patient 1.

## Discussion

In this study, we used a well-established viral dynamic model to show that early treatment with Paxlovid may preserve target cells which can then lead to viral rebound after the end of treatment. Additionally, the precise time therapy is initiated, within-host viral dynamic parameters, an individuals’ specific response to Paxlovid (e.g., variation in pharmacokinetic and pharmacodynamics parameters– not shown), and the timing of adaptive immunity play important roles in determining whether viral rebound occurs. These important factors vary from individual to individual and may explain why only some individuals show viral rebound after completing treatment with Paxlovid.

Using parameter values within the range of literature values (see *Supplementary Materials*), we showed that a standard viral dynamic model without an adaptive immune response can capture the viral load changes observed in patient 1 including viral rebound and clearance (Fig. 1). We then showed that the model can also describe the viral load data in patients 2 and 3 when the effect of adaptive immunity is added to the model (Fig. 1).

Interestingly, the three patients studied here were all vaccinated and boosted and nonetheless had breakthrough infections with Omicron subvariant BA1.20 (patient 1) or BA2.9 (patients 2 and 3) (8). Thus, while adaptive immune responses may have occurred, they were either too weak or too late to prevent infection. In fact, our model suggests that in patient 1 the response even after breakthrough infection continued to be too weak or too late to affect the observed viral dynamics, i.e., the predicted time for the onset of the response *t*\* is past the last data point at day 21. One could speculate that an adaptive response occurred and preserved additional target cells but there is no data to support this speculation. For this reason, we chose not to include adaptive immunity for patient 1.

The viral dynamics observed in patients 1-3 can be captured equally well by an immune response (IR) model, where the protective effects of innate immunity can preserve target cells by putting them into an antiviral state (see *Supplementary Materials*). Again, for patient 1 there is no support for including an adaptive response. From a biological perspective the IR model is attractive but it introduces additional parameters, which are difficult to estimate due to the lack of quantitative data about the innate response in these viral rebounders.

Shortly after treatment begins, Paxlovid reduces viral production, which halts the decline in target cells. This preservation effect leaves target cells at an approximately constant level over the course of treatment. Once the 5-day course of treatment is complete, if the level of preserved target cells is sufficient, it can allow the virus to rebound. Through simulations, we showed that there are three key factors that determines whether the level of preserved target cells after the completion of a 5-day treatment with Paxlovid is sufficient to support viral rebound.

First, the timing of treatment plays a crucial role. If treatment is initiated too early, before a time we denote *t_critical_*, a substantial number of target cells remains after the 5-day treatment and viral rebound is likely to occur (Fig. 1). After *t_critical_* too few target cells remain to support viral growth. Since viral growth switches to viral decay at the time of the viral peak in an untreated individual, this means *t_critical_*is the time the viral peak is reached. In more technical terms *t_critical_* corresponds to the time the reproductive number *R* = 1, so that on average each infected cells infects exactly one other cell, so that there is neither growth nor decay in the number of infected cells and level of viremia. According to a human challenge study (15), *t_critical_* should be ∼ 5 days after infection, or about 1 to 3 days after symptom onset. This suggests that delaying treatment may be a strategy to reduce the possibility of viral rebound. However, Paxlovid treatment accelerates viral clearance and hence potentially can reduce viral transmission. Thus, delaying treatment may have a detrimental effect on public health and deserves more study. In addition, delaying treatment could also have impact on the severity of disease in the high-risk patients for whom Paxlovid is recommended.

All viral dynamic parameters affect *R* and hence they affect *t_critical_*. Due to the differences in viral dynamic parameters from patient to patient, *t_critical_* is different for each patient, which means the best time to start treatment to obtain both a rapid viral decline and prevent viral rebound is likely to be different for each patient. However, in general, delaying treatment may decrease the probability of developing rebound (Figs. 3).

While adaptive B and T cell immune responses may have occurred too late to prevent infection, they could still have affected the level of preserved target cells. Thus, their timing and emergent speed may play a crucial role in the occurrence of viral rebound. Early treatment with Paxlovid reduces the viral load and number of infected cells, which may delay both the innate and adaptive immune responses and so may also contribute to the observed viral rebound.

In summary, our model suggests the occurrence of viral rebound following a complete course of Paxlovid may be due to incomplete virus clearance and the level of preserved target cells. Increasing the length of treatment from 5 to 10 days will continue to preserve target cells and thus may still allow viral rebound if viable virus is present at the end of treatment and sufficient adaptive immunity has not developed. Furthermore, starting an additional course of treatment one day after symptom rebound makes little difference in the viral clearance time. However, delaying initiation of treatment for a day or two after having symptoms or testing positive may have some benefit in reducing the possibility of rebound, but at the cost of allowing viral growth to continue and the possibility of increased viral transmission and detrimental effects on the course of disease severity. Clinical trials could evaluate these various potential treatment strategies. Lastly, rebound following antiviral treatments is not unique to Paxlovid (13). These findings may provide an explanation to rebound following other antiviral treatments besides Paxlovid.

## Methods

We use two models to study viral rebound, a standard viral dynamic model related to the one developed by Baccam et al. (16) to study acute influenza infections and an expanded version of that model that includes an innate response involving type I interferon (IFN), called the IR model. Both models make very similar predictions. Thus, we present the simpler model without an innate response in the main text while the model with IFN is discussed in the *Supplementary Materials*.

The model we use is described by the following set of ordinary differential equations:

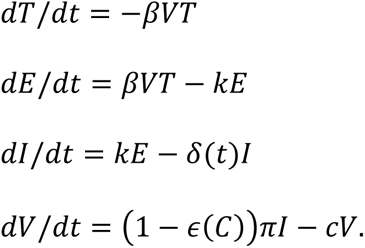

In this model, *T* is the number of target cells, *E* is the number of infected cells that have not yet started to produce virus, i.e., are in the eclipse phase, *I* is the number of productively infected cells, and *V* is the viral load. Target cells become infected with rate constant *β*. After being infected for an average time of 1/*k*, infected cells start producing virus at an adjusted rate *π* that accounts for sampling via a swab (14) and die at per capita rate *δ*, which we allow to be time dependent as described below. SARS-CoV-2 viruses are cleared at per capita rate *c*. The effectiveness of nirmatrelvir in blocking viral replication and subsequent production of virions is given by 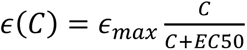, an E_max_ model where *C* is the concentration of nirmatrelvir, *EC*_50_ is the concentration at which the drug effectiveness is half-maximal and *∈*_max_is the maximum effectiveness. When *ε*(*C*) = 0 the drug has no effect and when *ε*(*C*) = 1 the drug is 100% effective at blocking virion production.

We include in this model (and the IR model in the *Supplementary Materials*) a possible adaptive immune response, since rebounds tend to occur late after infection, when the adaptive immune response is expected to be important. As in a previous study by Pawelek et al. (17), we added this response to the model starting at time *t*^∗^. We assume that the adaptive response increases exponentially at rate *σ* and causes an increase in the death rate of infected cells. This increased death rate could be due to the increasing presence of cytotoxic T cells or of viral-specific antibodies that bind to infected cells and cause their death by processes such as antibody-dependent cytotoxicity, antibody-dependent phagocytosis or complement-mediated death. To account for these phenomena, we use the following time-dependent infected cell death rate *δ*(*t*):

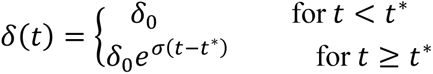

As it is impossible to know the number of viruses that initiated infection, we use a method suggested by Smith et al. (18) in which we assume the initiating virus is either cleared or rapidly infects cells. Thus, as initial conditions we use: *T*(0) = 8 × 10^7^ cells, *E*(0) = 1 cell, *I*(0) = 0, *V*(0) = 0, and *R*(0) = 0 as explained in Ke et al. (14). We note that the infection dynamics are relatively insensitive to increasing the initial number of infected cells to 10 (14).

### Pharmacokinetic model for Paxlovid

We assume the drug effectiveness *ε*(*C*) depends on the concentration of nirmatrelvir, *C*(*t*), according to an *E_max_* model with EC50 = 62 nM (1). Following a *single dose* of nirmatrelvir of 300 mg with 100 mg of ritonavir, the observed maximum nirmatrelvir concentration is 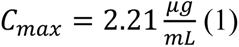. As nirmatrelvir has a molecular weight of 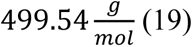 this value of *C_max_* can also be expressed as 4.4 × 10^3^ *nM*. The half-life of nirmatrelvir when taken with ritonavir is about 6 hours (1), which corresponds to an elimination rate of 2.8/day. Additionally, dosing twice-daily achieved steady-state on day 2 with approximately 2-fold accumulation (1). Using a simple multidose absorption-elimination model, the pharmacokinetics of nirmatrelvir is given by (20)

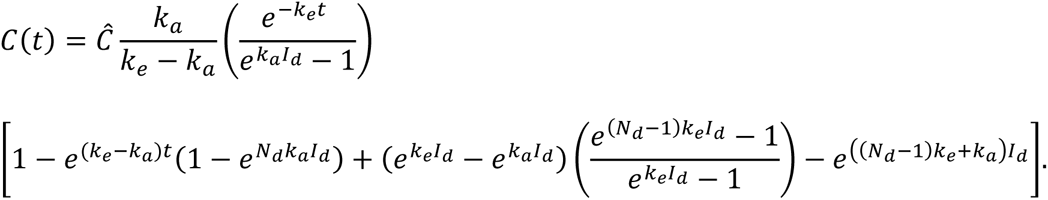

Here, *k_e_* is the elimination rate (2.8/day), *k_a_* is the absorption rate (17.5/day), *I_d_* is the dosing interval (1/2 day), 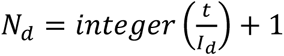 is the number of doses until time *t*, with the first dose at time 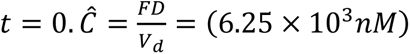, where *F* is the bioavailability of the drug, *D* is the mass of the drug administered in a dose (300 mg), and *V_d_* is the volume of distribution. Details on the implementation of the pharmacokinetic model and the parameter values used can be found in the *Supplemental Material*. With these assumptions, the drug effectiveness *ε*(*C*) hovers around 0.98 during treatment (Fig. S1) and then falls to zero rapidly after treatment stops (Fig. S2).

### Data fitting

For comparison between our model and data, we converted the PCR cycle threshold (Ct) values reported in Charness et al. (8) to log_10_ RNA copies (see *Supplemental Material*). Viral load data for patients 2 and 3, using a different PCR system, were kindly provided by David Ho (*Supplemental Material*). The duration from the time of infection to symptom onset is assumed to be 3 days for all three patients, which is midway between the 2-4 days reported in a human challenge study (15). The time after symptom onset when treatment started is reported in Charness et al. (8), as day 0, day 1, and day 2 for patients, 1, 2 and 3 respectively, but for patient two we used 12 hours, which is more accurate as reported in (21).

We fit the model to viral load data using the optimization function *fminsearch* (MATLAB 2021b), which uses the Nelder-Mead simplex direct search method, to minimize the difference:

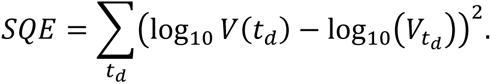

Here, *t_d_* is the time that corresponds to the available data *V_td_*.

## Data Availability

There are no original data underlying this work. Only previously published data were used for this study by Charness et al. (2022). The viral load values used in this study are given in the Supplementary Materials.
M. E. Charness et al., Rebound of SARS CoV 2 Infection after Nirmatrelvir Ritonavir Treatment. N. Engl. J. Med. 387, 1045-1047 (2022).

## Funding

This work was performed under the auspices of the US Dept. of Energy under contract 89233218CNA000001 and supported by

National Institutes of Health grant R01-OD011095 (ASP)

National Institutes of Health grant R01-AI116868 (RMR)

National Institutes of Health grant U54-HL143541-04 (RMR)

Los Alamos National Laboratory LDRD 20200743ER, 20200695ER, and 20210730ER.

## Author contributions

Conceptualization: ASP, RMR, TP

Methodology: ASP, RMR, TP

Investigation: ASP, RMR, TP

Visualization: TP

Funding acquisition: ASP, RMR

Project administration: ASP

Supervision: ASP, RMR

Writing – original draft: TP

Writing – review & editing: ASP, RMR, TP

## Data and materials availability

There are no original data underlying this work. Only previously published data were used for this study (8). The viral load values used in this study are given in the Supplementary Materials.

## Supplementary Materials

Supplementary Text

Figs. S1 to S10 Tables S1 to S2

References (*22–25*) are only cited in the Supplementary Materials

Data S1

---

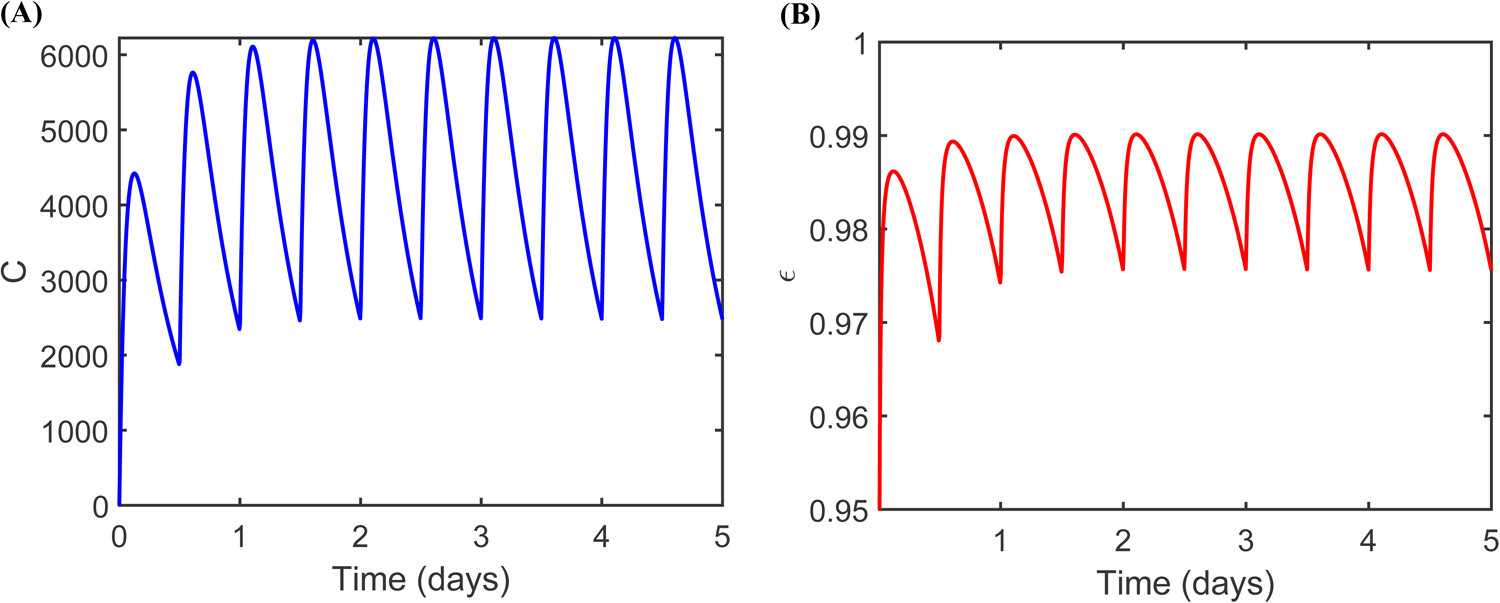

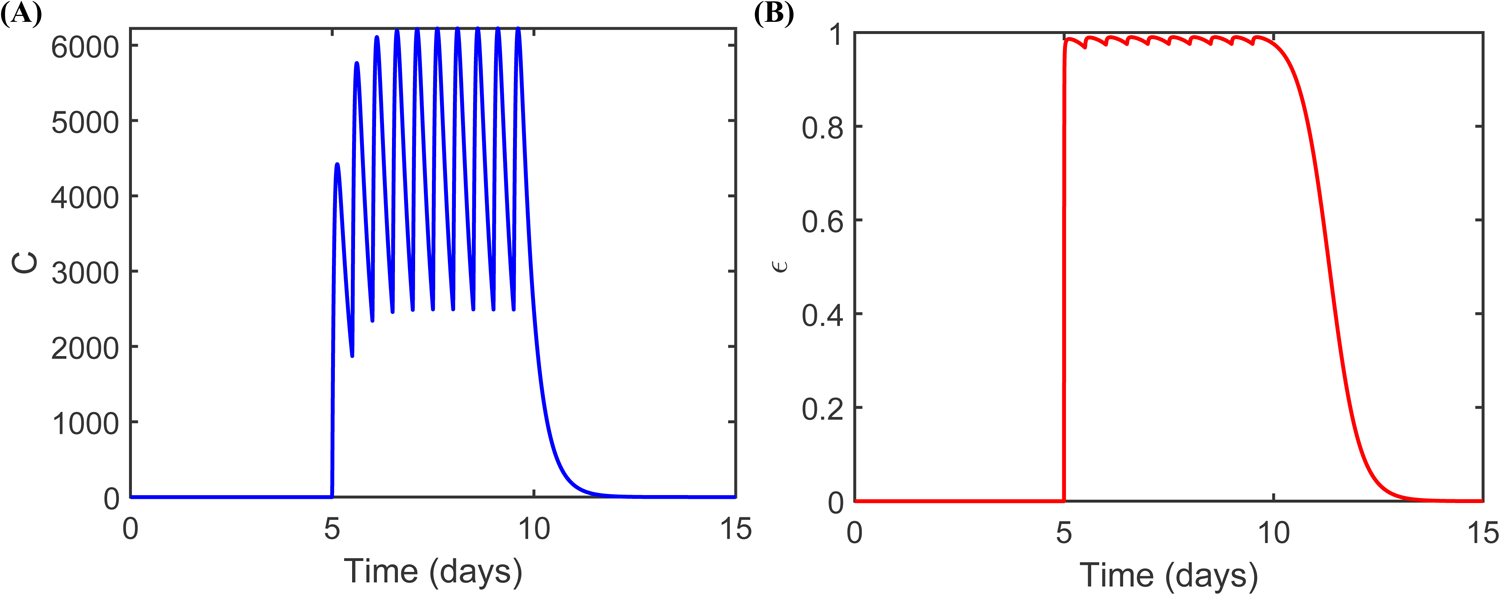

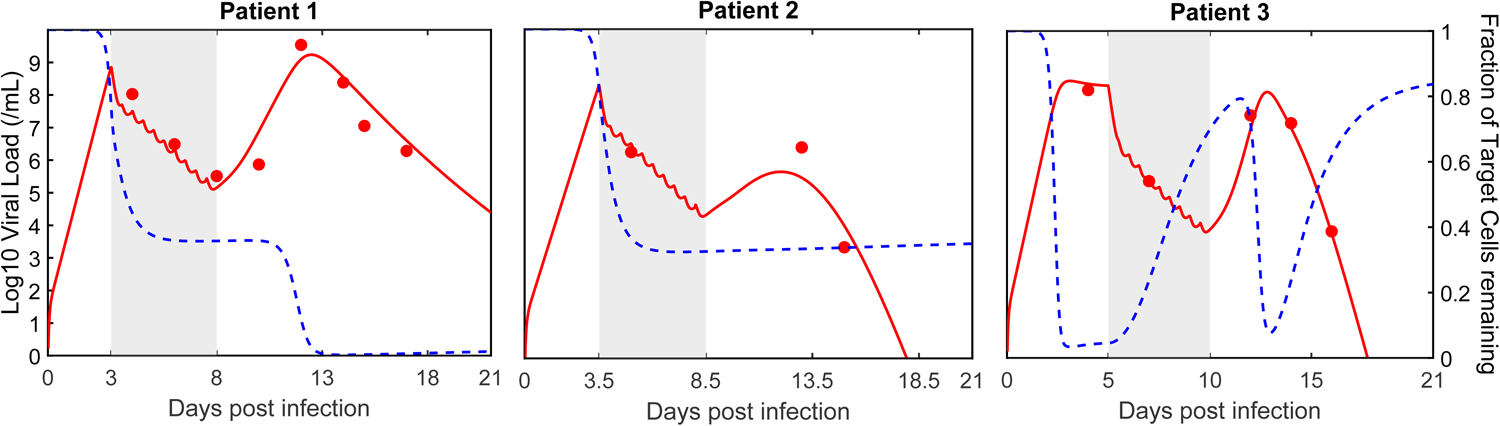

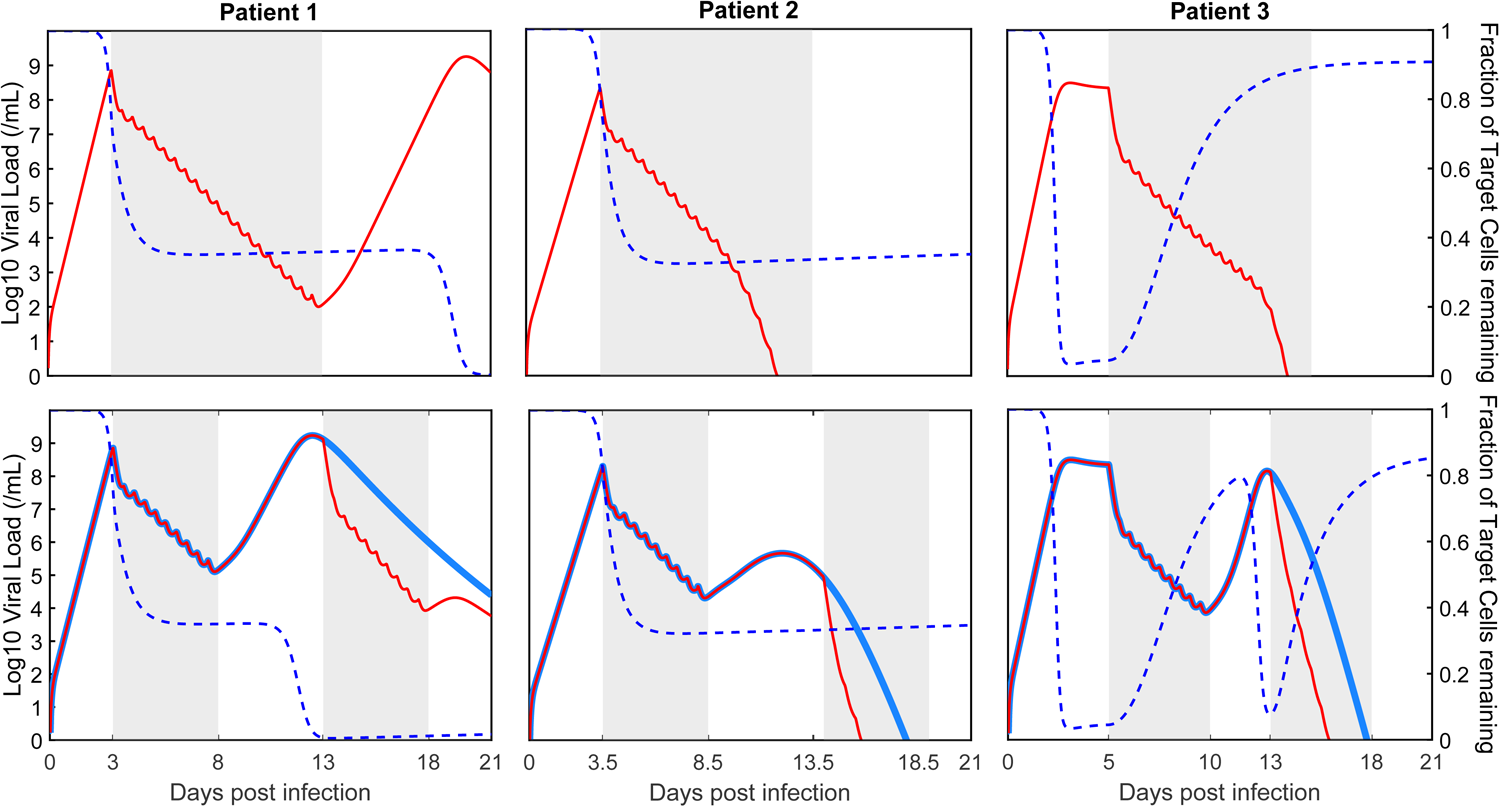

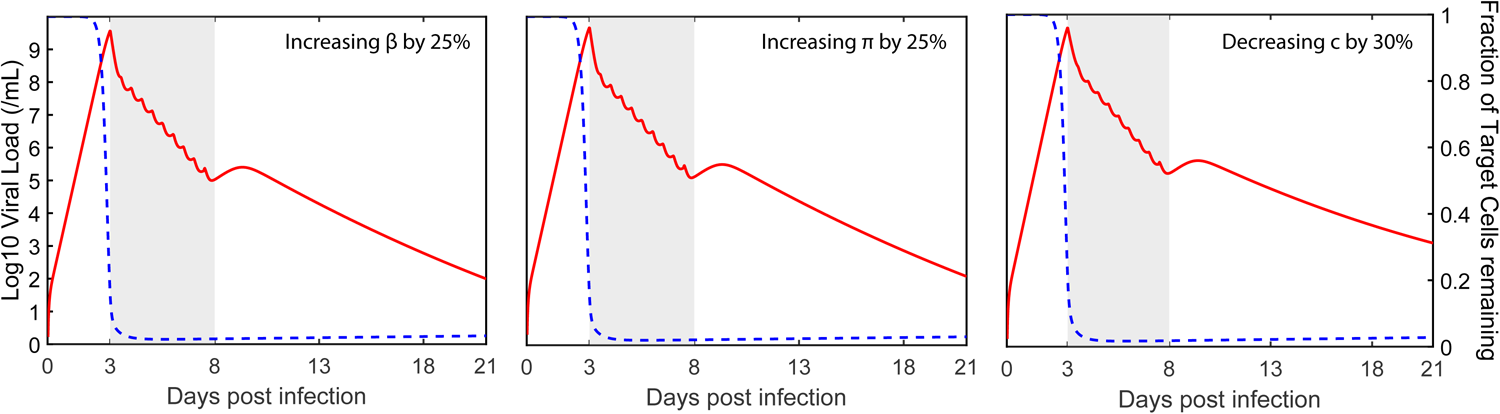

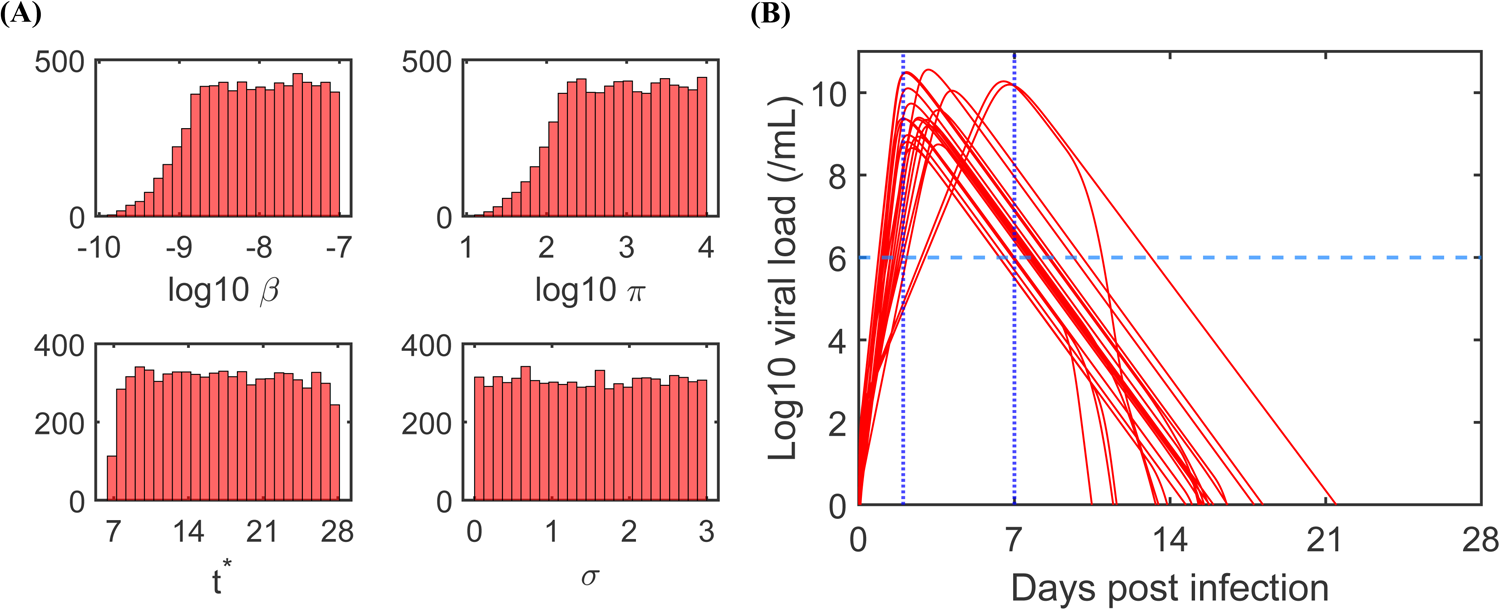

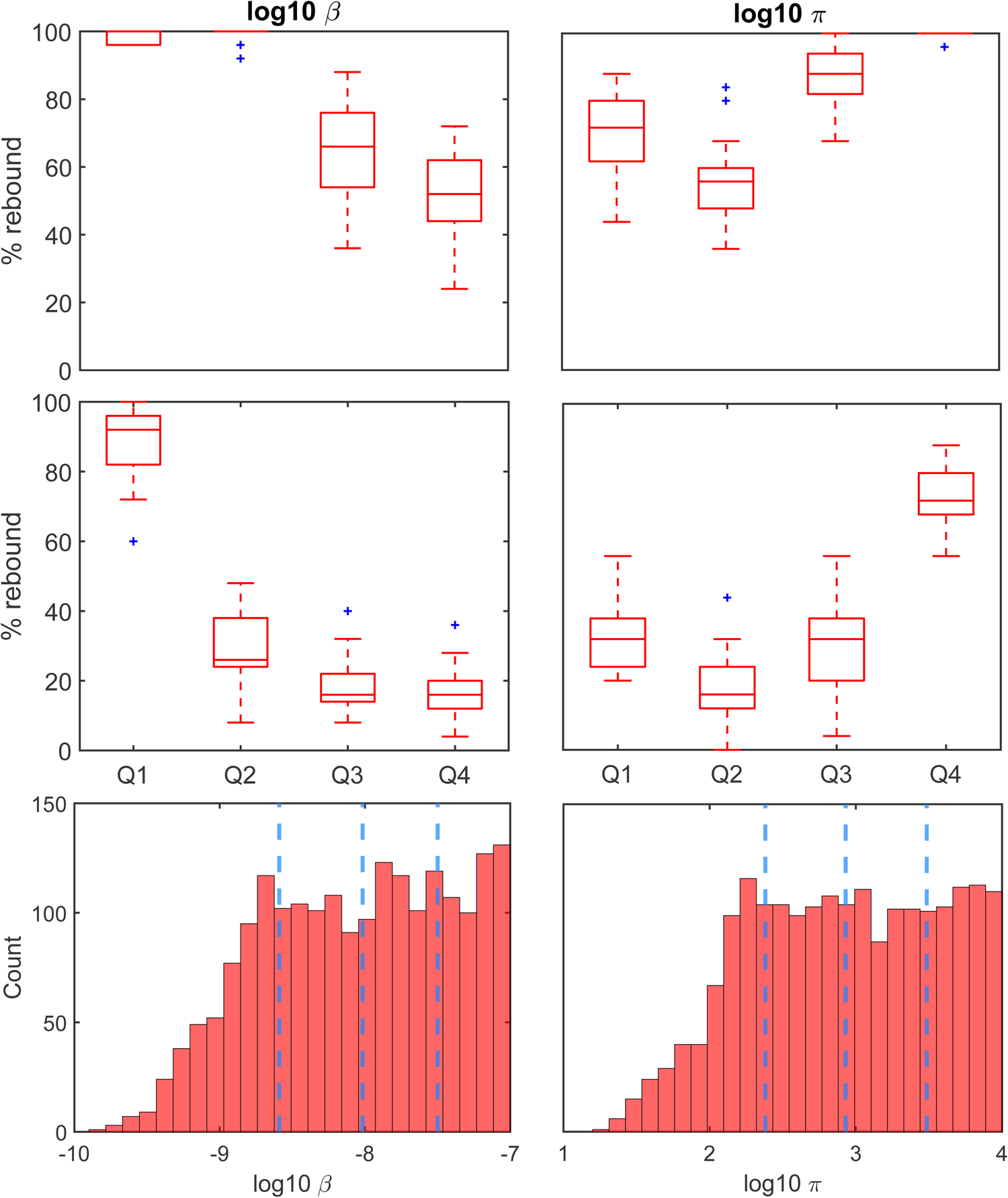

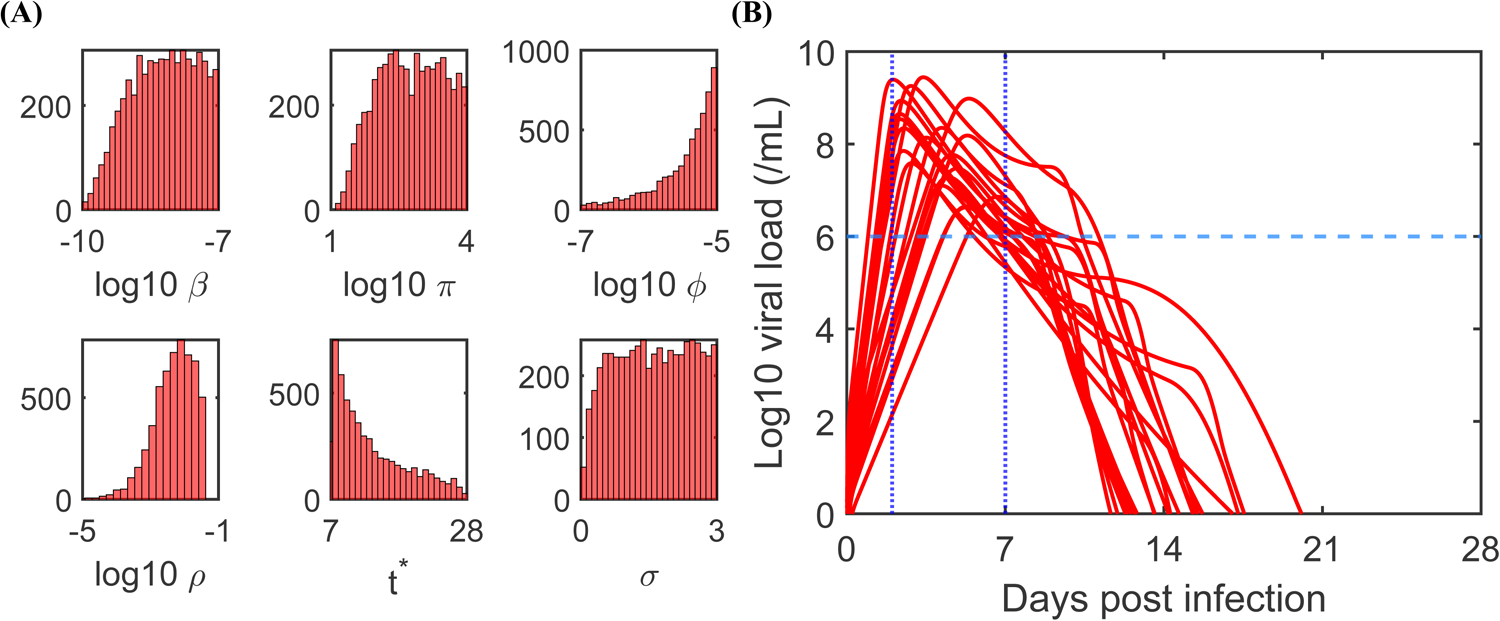

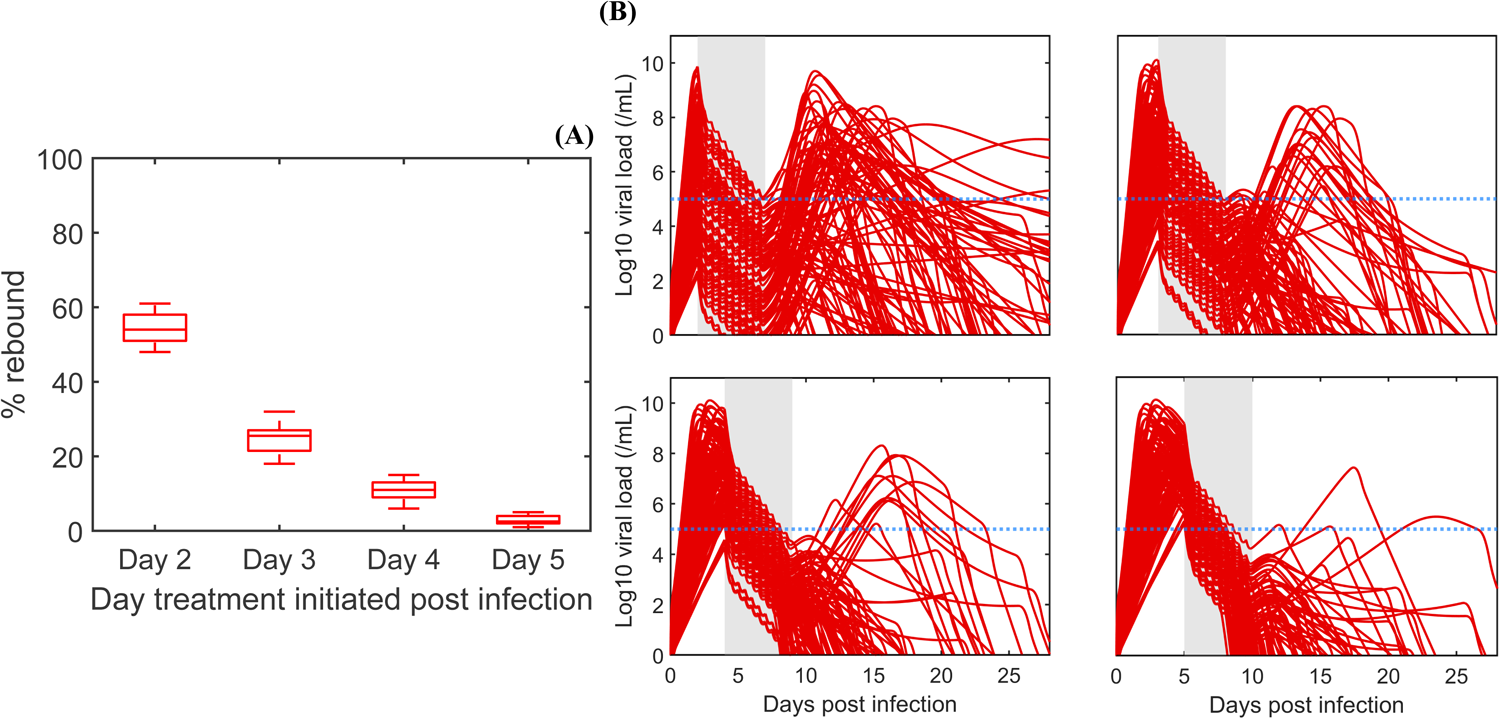

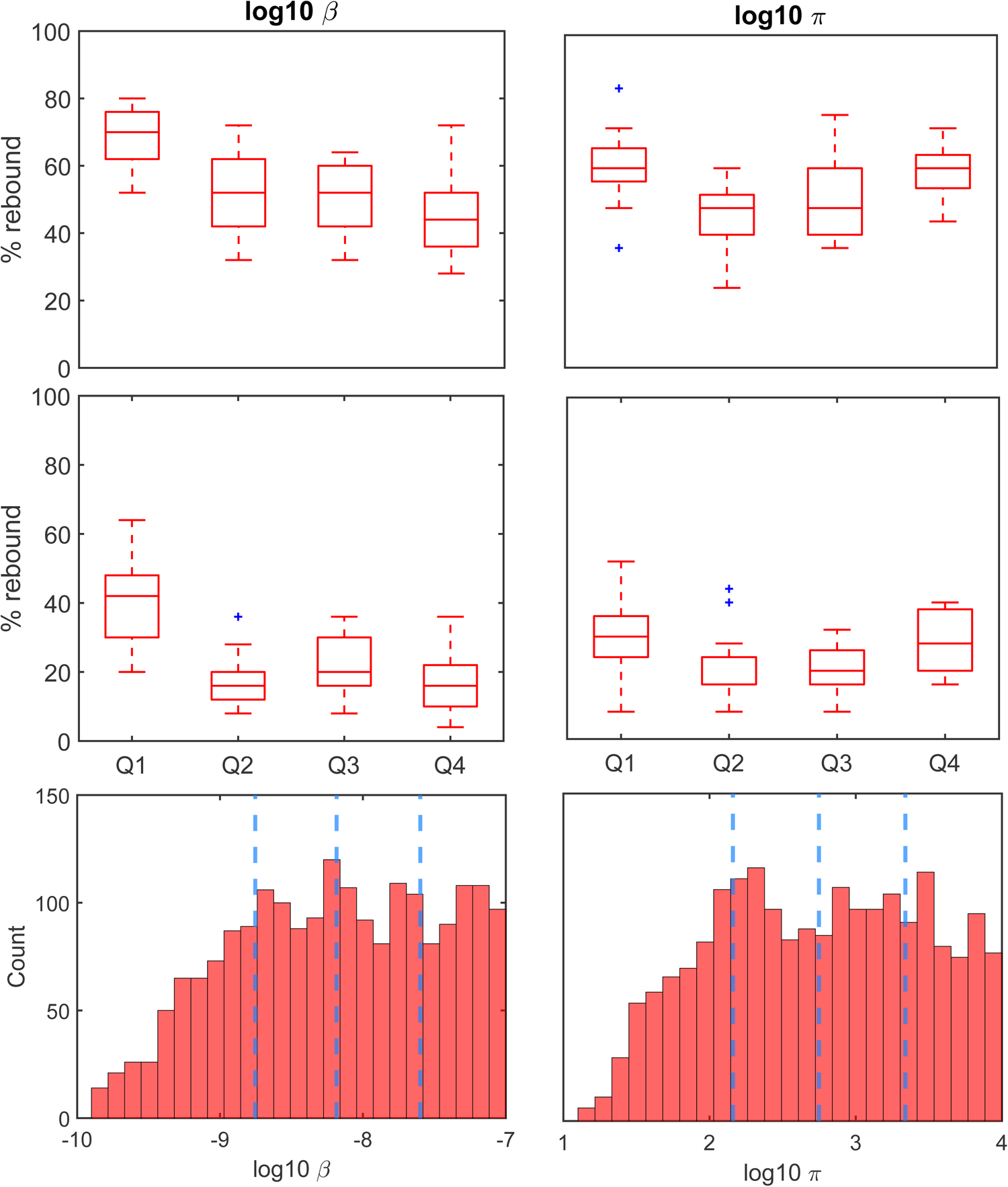

## References and Notes

1. U. S. Food and Drug Administration. Center for Drug Evaluation and Research. (2021). Fact Sheet for Healthcare Providers: Emergency Use Authorization for PAXLOVID. Washington, DC. https://www.fda.gov/media/155050/download. (Accessed 06/28/2022).

2. U. S. Food and Drug Administration. Center for Drug Evaluation and Research. FDA Updates on Paxlovid for Health Care Providers (2022). Washington, DC. https://www.fda.gov/drugs/news-events-human-drugs/fda-updates-paxlovid-health-care-providers. (Accessed 06/28/2022).

3. C. K. H. Wong et al., Real-world effectiveness of molnupiravir and nirmatrelvir plus ritonavir against mortality, hospitalization, and in-hospital outcomes among community-dwelling, ambulatory patients with confirmed SARS-CoV-2 infection during the omicron wave in Hong Kong: an observational study. The Lancet 400,1213–1222 (2022).

4. J. Hammond et al., Oral nirmatrelvir for high-risk, non-hospitalized adults with Covid-19. N. Engl. J. Med. 386, 1397–1408 (2022).

5. R. Arbel et al., Nirmatrelvir use and severe COVID-19 outcomes during the Omicron Surge. N. Engl. J. Med. 387, 790–798 (2022).

6. Centers for Disease Control and Prevention. COVID-19 rebound after Paxlovid treatment. U.S. Department of Health and Human Services (2022). https://emergency.cdc.gov/han/2022/pdf/CDC_HAN_467.pdf. (Accessed 06/28/2022)

7. J. Boucau et al., Characterization of Virologic Rebound Following Nirmatrelvir-Ritonavir Treatment for Coronavirus Disease 2019 (COVID-19). Clin. Infect. Dis. Jun 23, ciac512 (2022).

8. M. E. Charness et al., Rebound of SARS-CoV-2 Infection after Nirmatrelvir–Ritonavir Treatment. N. Engl. J. Med. 387, 1045–1047 (2022).

9. H. Soares et al., Viral load rebound in placebo and nirmatrelvir-ritonavir treated COVID-19 patients is not associated with recurrence of severe disease or mutations. Research Square [Preprint] (2022). https://doi.org/10.21203/rs.3.rs-1720472/v2. (Accessed 06/28/2022).

10. A. F. Carlin et al., Virologic and Immunologic Characterization of Coronavirus Disease 2019 Recrudescence After Nirmatrelvir/Ritonavir Treatment. Clin. Infect. Dis. Jun 20, ciac496 (2022).

11. N. Ranganath et al., Rebound phenomenon after nirmatrelvir/ritonavir treatment of coronavirus disease-2019 in high-risk persons. Clin. Infect. Dis., ciac481 (2022).

12. E. Y. Dai et al., Viral kinetics of Severe Acute Respiratory Syndrome Coronavirus 2 (SARS-CoV-2) Omicron infection in mRNA-vaccinated individuals treated and not treated with nirmatrelvir-ritonavir. medRxiv [Preprint] (2022). https://doi.org/10.1101/2022.08.04.22278378. (Accessed 09/03/2022).

13. L. Wang et al., COVID-19 rebound after Paxlovid and Molnupiravir during January-June 2022. medRxiv [Preprint] (2022). https://doi.org/10.1101/2022.06.21.22276724. (Accessed 12/23/2022).

14. R. Ke, C. Zitzmann, D. D. Ho, R. M. Ribeiro, A. S. Perelson, In vivo kinetics of SARS-CoV-2 infection and its relationship with a person’s infectiousness. Proc. Natl. Acad. Sci. 118, e2111477118 (2021).

15. B. Killingley et al., Safety, tolerability and viral kinetics during SARS-CoV-2 human challenge in young adults. Nat. Med. 28, 1031–1041 (2022).

16. P. Baccam, C. Beauchemin, C. A. Macken, F. G. Hayden, A. S. Perelson, Kinetics of Influenza A virus infection in humans. J. Virol. 80, 7590–7599 (2006).

17. K. A. Pawelek et al., Modeling within-host dynamics of influenza virus infection including immune responses. PLoS Comput. Biol. 8, e1002588 (2012).

18. A.P. Smith, D. J. Moquin, V. Bernhauerova, A. M. Smith, Influenza virus infection model with density dependence supports biphasic viral decay. Front. Microbiol. 9 1554 (2018).

19. National Center for Biotechnology Information. PubChem Compound Summary for CID 155903259, nirmatrelvir (2022). https://pubchem.ncbi.nlm.nih.gov/compound/155903259. (Accessed 06/28/2022).

20. N. M. Dixit, A. S. Perelson, Complex patterns of viral load decay under antiretroviral therapy: influence of pharmacokinetics and intracellular delay. J. Theor. Biol. 226, 95–109 (2004).

21. R. Rubin, From positive to negative to positive again—the mystery of why COVID-19 rebounds in some patients who take Paxlovid. JAMA 327.24: 2380–2382 (2022).

22. T. C. Jones et al., Estimating infectiousness throughout SARS-CoV-2 infection course. Science 373.6551: eabi5273 (2021).

23. E. A. Hernandez-Vargas, J. X. Velasco-Hernandez, In-host mathematical modelling of COVID-19 in humans. Annu. Rev. Control 50: 448–456 (2020).

24. N. Néant et al., Modeling SARS-CoV-2 viral kinetics and association with mortality in hospitalized patients from the French COVID cohort. Proc. Natl. Acad. Sci. U.S.A. 118.8: e2017962118 (2021).

25. A. Goyal, E. F. Cardozo-Ojeda, J. T. Schiffer, Potency and timing of antiviral therapy as determinants of duration of SARS-CoV-2 shedding and intensity of inflammatory response. Sci. Adv. 6.47: eabc7112 (2020).

