## Supplementary Material for "An explanation for SARS-CoV-2 rebound after Paxlovid treatment"

**The PDF file includes:**

Supplementary Text

Figs. S1 to S10

Tables S1 to S2

Data S1

**Conversion of Ct values to VL data**

We used a conversion formula that was developed specifically for the Roche Cobas 6800 system (22), the system used by the Charness lab to obtain data for patient 1 (8).

$$Log10\ Viral\ Load = 15.043 - (Ct \times 0.296)$$

**Nirmatrelvir concentration and drug effectiveness dynamics**

The concentration of nirmatrelvir is modeled based on a two-compartment model with first-order absorption rate $k_a$ and first-order elimination rate $k_e$ (20).

$$C(t) = \hat{C}\frac{k_a}{k_e-k_a}\left(\frac{e^{-k_e t}}{e^{k_a I_d}-1}\right)\left[1 - e^{(k_e-k_a)t}(1-e^{N_d k_a I_d}) + (e^{k_e I_d} - e^{k_a I_d})\left(\frac{e^{(N_d-1)k_e I_d}-1}{e^{k_e I_d}-1}\right) - e^{((N_d-1)k_e+k_a)I_d}\right],$$

where $I_d$ is the dosing interval (1/2 day), $N_d = integer\left(\frac{t}{I_d}\right) + 1$ is the number of doses until time $t$.

Based on pharmacokinetics of nirmatrelvir taken with ritonavir (1), the elimination rate $k_e$ is 2.8/day. Finding the maximum concentration from a single dose gives

$$C_{max} = \hat{C}\left(\frac{k_e}{k_a}\right)^{\frac{k_e}{k_a-k_e}} = 4.42 \times 10^3\ nM$$

which according to (1) takes place at $t_{max} = \frac{\ln\left(\frac{k_a}{k_e}\right)}{k_a-k_e} = 3\ hours = \frac{1}{8}\ day$. Solving for $k_a$ from the expression for $t_{max}$ gives $k_a = 17.5$/day. Then, solving for $\hat{C}$ in the expression for $C_{max}$ gives:

$$\hat{C} = \frac{C_{max}}{\frac{k_e}{k_a}^{\frac{k_e}{k_a-k_e}}} = 6.25 \times 10^3\ nM.$$

The drug effectiveness is given by $\epsilon(C) = \epsilon_{max}\left(\frac{C}{C+EC50}\right)$, where EC50= 62nM (3) and we assume the maximum drug effectiveness $\epsilon_{max} = 1$ for all of our simulations and model fits to data. Graphs of $C(t)$ and $\epsilon(C(t))$ for the parameters given above are provided in Figure S1.

Figure S2 shows a simulation for a 5-day course of Paxlovid starting on day 5 and ending on day 10. Note that unlike Figure S1, Figure S2 shows the drug elimination phase after the end of the treatment.

In both Figures S1 and S2, the drug concentration reaches a stable oscillatory level, above 50 times EC50, around day 2, which is consistent with the observation that drug concentration stabilizes after two days (1) and leads to a quasi-constant high efficacy of ~0.98.

**A note on model parameters**

All the parameter values used for the model simulations are within previously estimated ranges (14, 17). In particular, the average products of $\beta\pi$ and $\beta T_0$ in these patients are on the order of $10^{-6}$ and $10^{-1}$, respectively. The reported ranges for $\beta\pi$ and $\beta T_0$ in the literature are on the order of $10^{-7} - 10^{-5}$ and $10^{-1} - 10^{1}$, respectively (23-25).

**Immune response (IR) model**

We extended the viral dynamic model to incorporate the effect of innate and adaptive immunity.

$$T' = -\beta VT - \phi IT + \rho R$$

$$R' = \phi IT - \rho R$$

$$E' = \beta VT - kE$$

$$I' = kE - \delta(t)I$$

$$V' = \big(1 - \epsilon(C)\big)\pi I - cV$$

The innate immune response is modeled as in Ke et al. (14) where it is assumed that the level of type-I interferon is proportional to the number of infected cells, $I$, and interferon puts target cells in an antiviral state that is refractory to infection at rate $\phi$. The number of cells refractory to infection is denoted $R$. Refractory cells lose their protection and become susceptible to infection at a rate $\rho$. Adaptive immunity is modeled as described in the main text.

For the IR model, we used the following initial values: $T(0) = 8 \times 10^7$ cells, $E(0) = 1$ cell, $I(0) = 0$, $V(0) = 0$, and $R(0) = 0$, as in Ke et al. (4).

**The IR model fit to VL data**

The IR model can fit the data and describe the observed viral rebound well for all three patients (Fig. S3). Furthermore, the IR model provides a significant improvement in the fit to patient 3 compared to the standard viral dynamic model (Table S1). The IR model gives lower fitting error in patient 1; however, due to two additional fitting parameters, the standard model is preferred in terms of AIC. For patient 2, it is possible for the IR model to give as good of a fit as the standard model (by setting $\phi = \rho = 0$). However, this would defeat the purpose of testing an alternative model. Thus, the initial guesses for those two parameters are chosen close to literature values instead.

Similar to the standard model, best-fit parameter values for the IR model are within previously estimated ranges by Ke et al. (14) and Pawelek et al. (17). In fact, $c$, $\delta$ are the same in both models, and $\beta, \pi$ are in similar ranges. We compare the model fits using AIC scores. The standard model has a lowest AIC score in patients 1 and 2, while the IR model has the lowest score for patient 3 as well as a better fit based on SSE (Table S1).

In Fig. S3, shortly after the treatment stops, the number of target cells remains at a plateau or starts to increase. For all three patients the level of preserved target cells is sufficient to allow the virus to rebound.

The results from the 10-day hypothetical treatment course with Paxlovid for the IR model is similar to those from the standard model (Fig. S4). The 10-day treatment course was able to suppress the viral rebound in patients 2 and 3 regardless of the high level of preserved target cells. On the other hand, patient 1 still shows rebound. The viral rebound is a direct consequence of the lack of adaptive immunity in the model for patient 1.

As in the standard model, increasing $\beta$ or $\pi$, or decreasing $c$, has an impact on the number of available target cells at the start of treatment (and end of treatment) and hence on the occurrence of viral rebound. Figure S5 shows that increasing either of $\beta$ or $\pi$ by 25% (or decreasing $c$ by 30%) eliminates viral rebound.

**Probability of rebound following treatment with Paxlovid in a cohort of *in silico* patients**

To quantify the chance of viral rebound after a 5-day course of treatment with Paxlovid, we simulate a cohort of *in silico* patients for each model (standard and IR). We used the following selection criteria to construct a cohort of *in silico* patients without treatment: (1) The viral load must peak above $10^6$ copies per mL; (2) The peak must be reached between day 2 and day 7; (3) The viral load must decline below $10^2$ copies per mL by day 28. This algorithm is akin to a rejection algorithm, where we sample parameters from a predetermined uniform distribution and only accept parameter sets that satisfy conditions (1) – (3). The ranges of uniform distributions that we sampled from are presented in Table S2. The ranges for $\beta$, $\pi$, $\phi$, $\rho$ were chosen to contain the values reported in (14). The range for $\sigma$ was chosen to cover the reported range in (17). The lower bound for $t^*$ was chosen ad-hoc under the assumption that the emergence of the adaptive immune response takes about one week post infection. The upper bound for $t^*$ was limited to 28 days due to the simulation duration of 28 days. Then, we apply Paxlovid to the cohort of in silico patients that obey the criteria and examine the percentage of *in silico* patients that rebound.

***Standard viral dynamic model with an adaptive immune response***

We obtained 7,638 admissible parameter sets that fulfil criteria 1-3 above, from 100,000 simulation runs. The distribution of the parameters for the admissible parameter sets and 20 samples of the accepted trajectories are shown in Fig. S6. We used these admissible parameter sets to simulate treatment starting at different days and calculates the probability of rebound as described in the main text. Briefly, we chose randomly 100 of these in silico patients and calculated the fraction of rebound out of these. We repeated this process 20 times and plot the results over the 20 trials as boxplots in Figure 3.

We also generalized the analyses of the effect of $\beta$ and $\pi$ on the probability of rebound (extension of Fig. 4 – main text). In Fig. S7, we show the percentage of *in silico* patients that rebound (the same ones used to generate Fig. 3A) stratified based on the values of $\beta$ and $\pi$ binned into quartiles

Q1-Q4 of their $\log_{10}$ values. We observe that higher infection rates $\beta$ correspond to a lower percentage of rebound when treatment was given 2- or 3-days post infection. On the other hand, a moderate increase in the viral production rate $\pi$ (from Q1 to Q2) corresponds to a decrease in the percentage of rebound, while very high viral production rates (Q3 and Q4) associate with higher percentage of rebound. This is possibly because a small increase in the rate of viral production may lead to faster depletion of target cells, which limits the possibility of rebound. But if the viral production rate is too high, it may allow the virus to rebound to an observable level even with a reduced number of target cells and a small number of the infected cells that have yet to be cleared by the end of treatment. Additionally, delaying treatment also decreases the percentage of rebound regardless of the stratified group, as shown in the main text.

***Immune response (IR) model***

We obtained 5,604 admissible parameter sets fulfilling criteria 1-3 above, from 500,000 simulation runs. The distribution of the parameters for the admissible parameter sets and 20 samples of the accepted trajectories are shown in Fig. S6.

Like our study of the effect of delaying treatment with the standard viral dynamics model in the main text, we simulated a trial with 100 *in silico* treated patients and assessed what percentage exhibited rebound, which was defined as the viral load returning above $10^5$ RNA copies per mL. We repeated this trial protocol 20 times each for treatment starting at days 2, 3, 4 and 5. In Fig. S8A, we present the boxplots of the percentage of rebounds obtained from these 20 trials. Examples from 100 viral trajectories for each treatment initiation time are provided in Fig. S8B.

Based on these *in silico* trial results, the model predicts that the probability of rebound decreases quickly with increasing delay in Paxlovid initiation, similar to our results with the standard model.

We also generalize the effect of varying viral dynamic parameters ($\beta$ and $\pi$) and immune parameters ($\phi$ and $\rho$, not shown) on the probability of rebound in the IR model (similar to Fig. S7). Fig. S10 shows the percentage of rebound stratified by the logarithm of values of $\beta$ and $\pi$. We noted a similar trend as observed in Fig. S7. Increasing $\beta$ generally corresponds to a decrease in the percentage of rebound. On the other hand, a small increase in $\pi$ (Q1 to Q2) tends to decrease the percentage of rebound, but large increase in $\pi$ slightly increase the percentage of rebound. Finally, regardless of the values of $\beta$ and $\pi$, the IR model also predicts that delaying treatment corresponds to a decrease in the percentage of rebound.

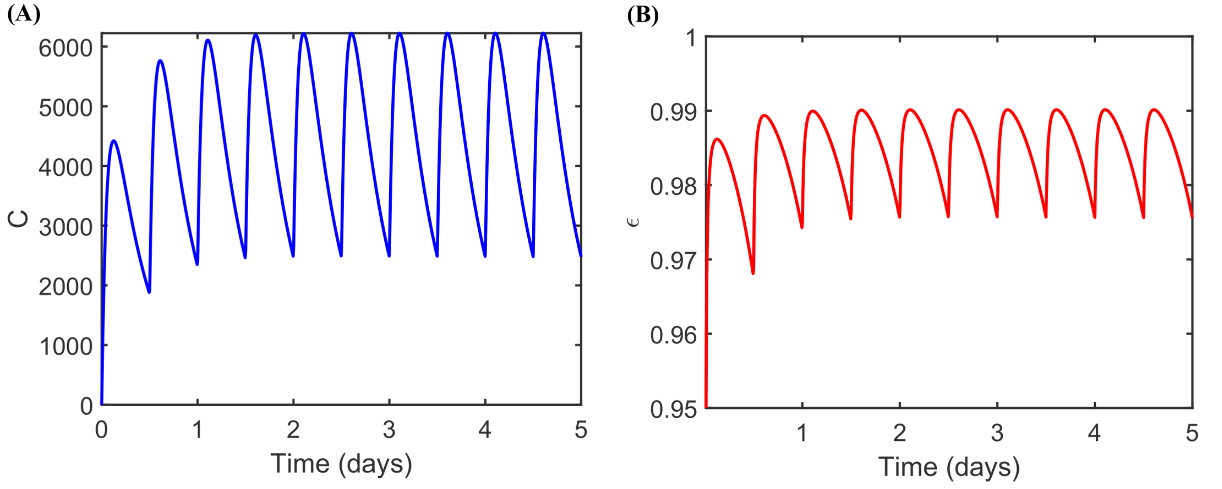


**Fig. S1.** Nirmatrelvir concentration over time based on the 5 day/10 dose schedule and the given kinetic parameters. (A) Nirmatrelvir concentration (nM). (B) The drug effectiveness over time based on nirmatrelvir concentration and an Emax model.

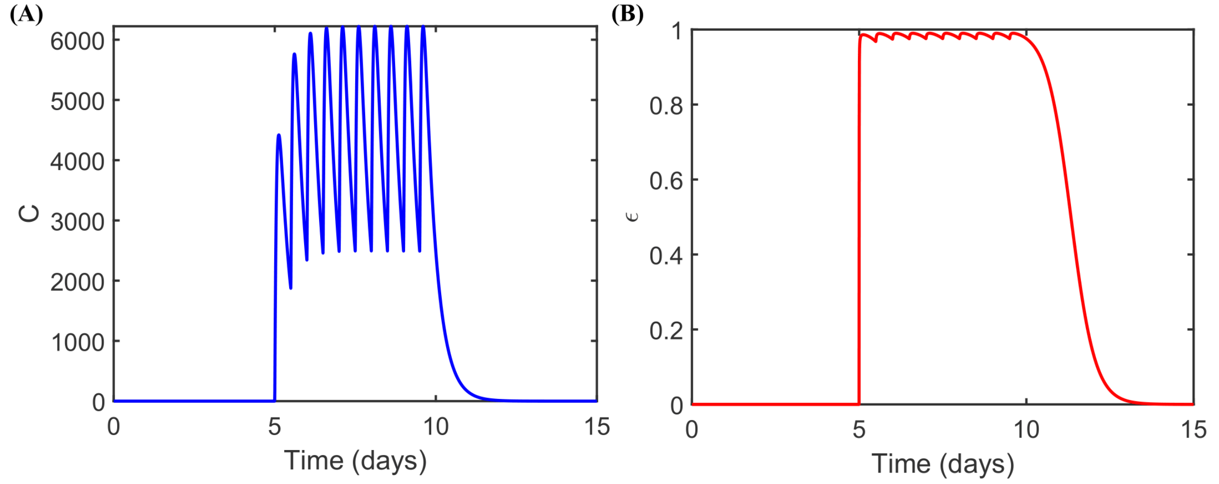


**Fig. S2.** Simulation of drug concentration (nM) and effectiveness over the 5 day/10 dose schedule starting on day 5 (with the last dose taken on day 9.5) with the given kinetic parameters. (A) Nirmatrelvir concentration (nM). (B) The drug effectiveness over time based on nirmatrelvir concentration and an Emax model.

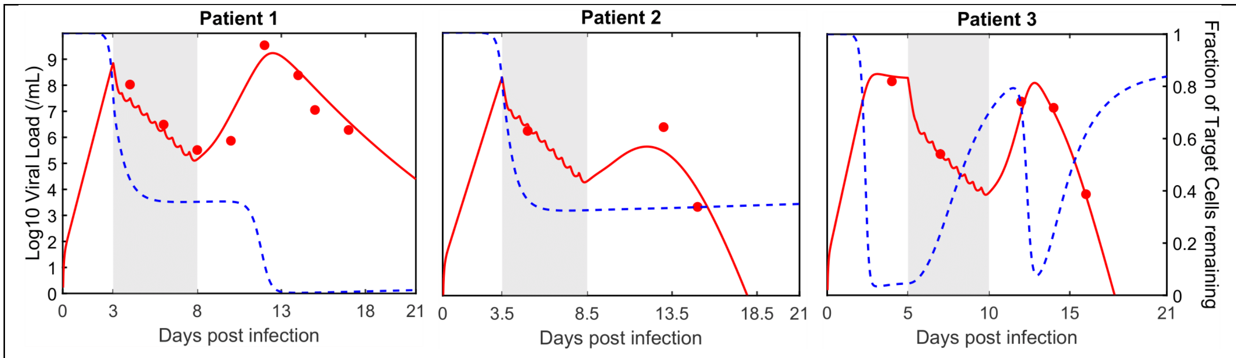


**Fig. S3.** Best fit of the IR model to the data. Solid red curves are the model simulated viral loads using the best-fit parameter values. Filled circles are the viral load data. The shaded area is the duration of treatment. The dashed blue curve is the fraction of target cells remaining. The IR model matches all three patients' data reasonably well. The following parameter values are shared across all three patients in the IR model: $\delta = 1.7\ \text{day}^{-1}, c = 10\ \text{day}^{-1}$, and $\sigma = 2.8\ \text{day}^{-1}$. For patient 1, $\beta = 1.70 \times 10^{-9}$ mL RNA copies$^{-1}$day$^{-1}$, $\pi = 2.11 \times 10^{3}$ RNA copies mL$^{-1}$day$^{-1}$, $\phi = 4.10 \times 10^{-8}$cells$^{-1}$day$^{-1}$, $\rho = 0.005\ \text{day}^{-1}$ and treatment runs from day 3 to 8. Similar to the result for the standard model, adaptive immunity is not needed for patient 1 in the IR model. For patient 2, $\beta = 2.81 \times 10^{-9}$ mL RNA copies$^{-1}$day$^{-1}$, $\pi = 9.28 \times 10^{2}$ RNA copies mL$^{-1}$day$^{-1}$, $\phi = 1.19 \times 10^{-7}$cells$^{-1}$day$^{-1}$, $\rho = 0.004\ \text{day}^{-1}, t^{*} = 8.61$ days, $\sigma = 0.40\ \text{day}^{-1}$ and treatment runs from day 3.5 to 8.5. For patient 3, $\beta = 2.21 \times 10^{-9}$ mL RNA copies$^{-1}$day$^{-1}$, $\pi = 2.00 \times 10^{3}$ RNA copies mL$^{-1}$day$^{-1}$, $\phi = 8.75 \times 10^{-6}$cells$^{-1}$day$^{-1}$, $\rho = 0.51\ \text{day}^{-1}, t^{*} = 12.00$ days, $\sigma = 1.27\ \text{day}^{-1}$ and treatment runs from day 5 to 10.

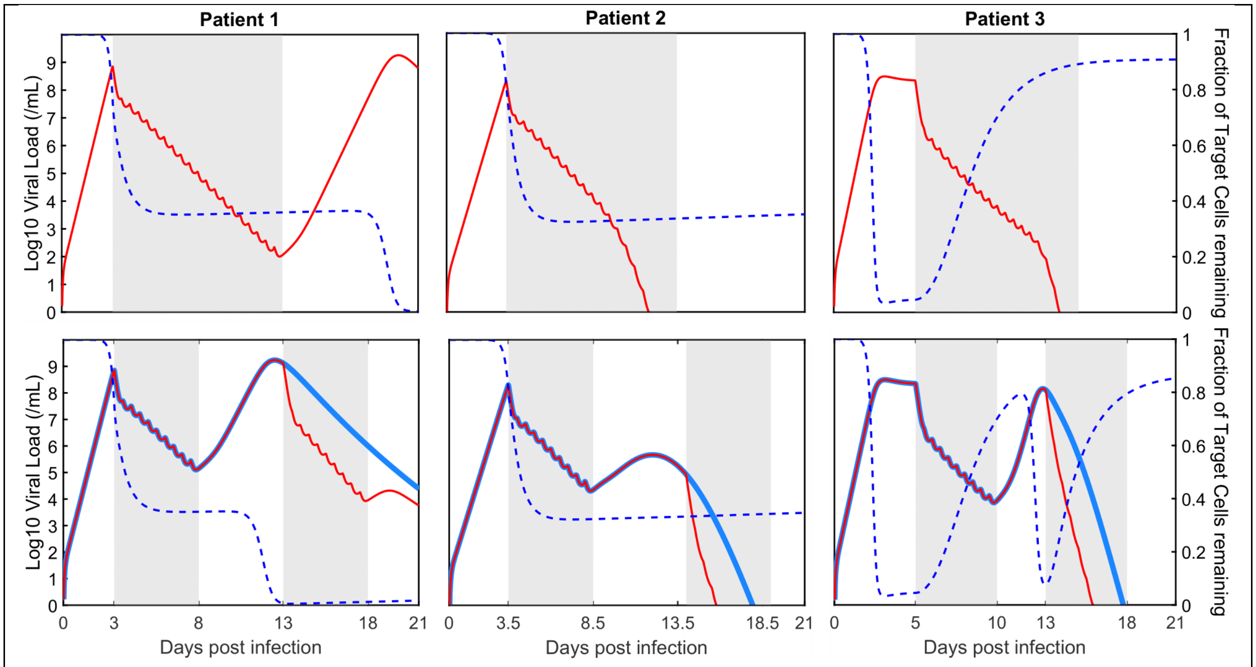


**Fig. S4.** IR model simulation with a 10-day hypothetical treatment: (A) 10 consecutive days; (B) one additional Paxlovid course one day after symptom rebound. Solid red curves are the model simulated viral load. The dashed blue curve is the model predicted number of target cells under the hypothetical treatment. The solid blue curves in (B) are the predicted viral trajectory with the standard five-day course of Paxlovid reproduced from Fig. S3. The shaded area is the duration of treatment. Giving a second course of treatment shortens the time to viral clearance by a few days for patients 2 and 3 and hardly has any effect for patient 1 where the solid red and green curves approach each other at longer times (not shown).

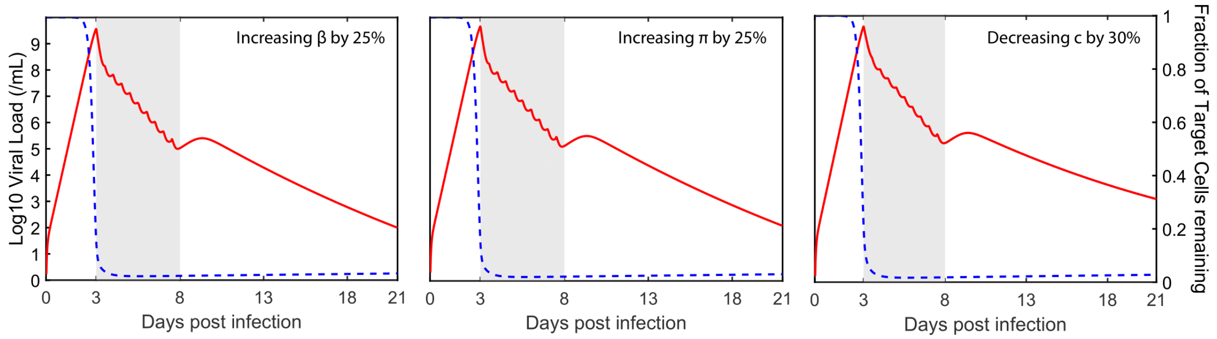


**Fig. S5.** IR model simulation with variation in values of $\beta$, $\pi$, and $c$ for patient 1. Solid red curves are the model simulated viral load. The blue dashed curve is the number of target cells. The shaded area is the duration of treatment. In the left, center and right panels, the model is simulated with $\beta$ 25% higher, $\pi$ 25% higher, and $c$ 30% lower than in the original simulations in Figure S3 for patient 1, respectively.

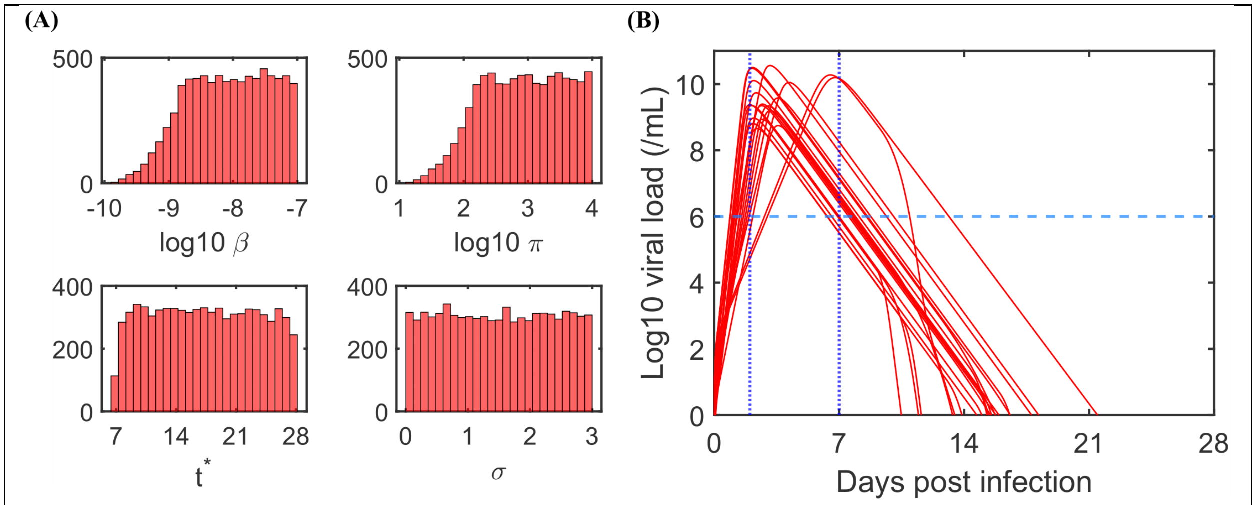


**Fig. S6.** (A) Distribution of accepted parameters for the standard model with an adaptive immune response. (B) 20 samples of accepted trajectories. The vertical blue dotted lines and the horizontal blue dashed line are the constraints on the viral peak.

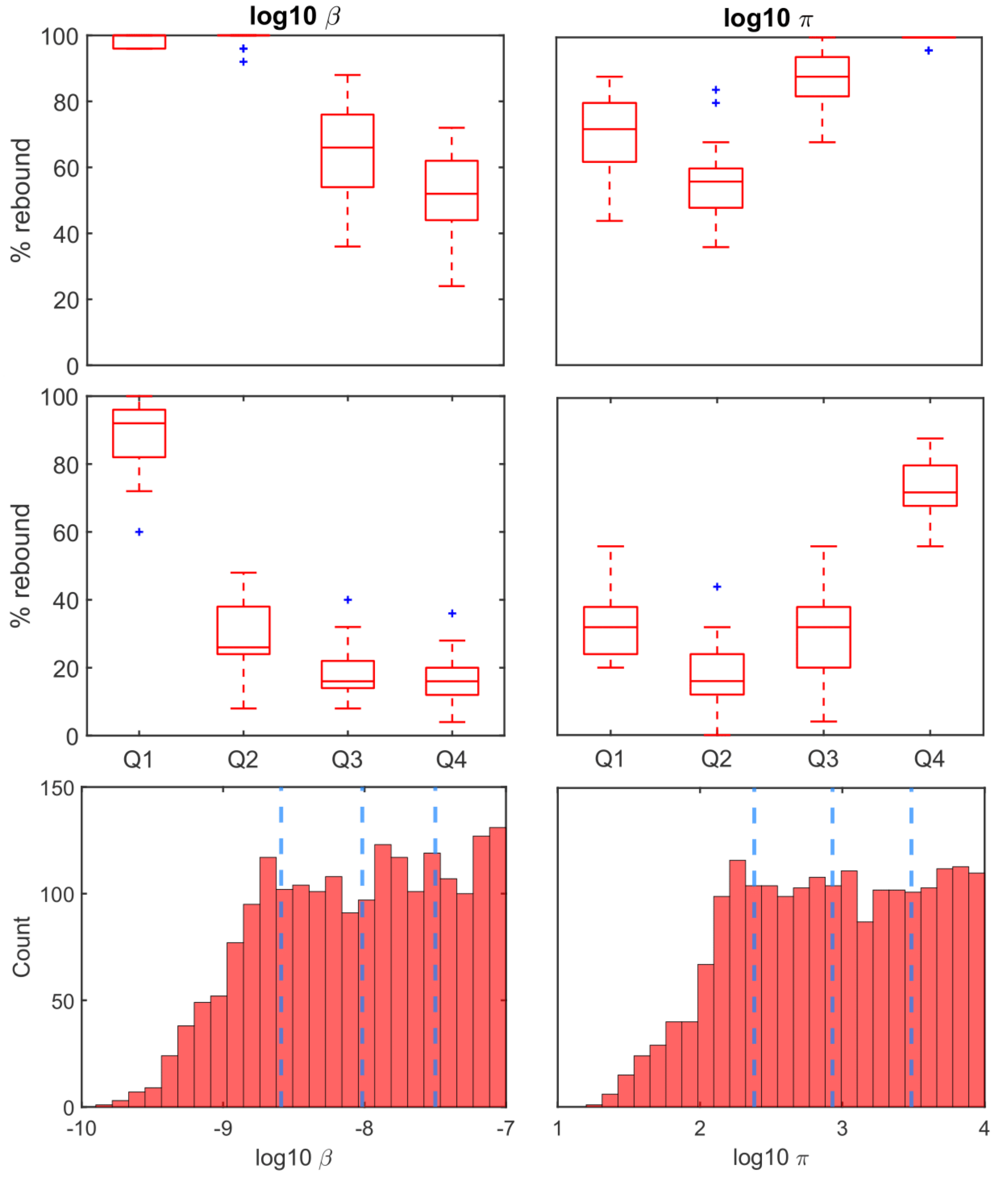


**Fig. S7.** Percentage of predicted *in silico* patients with rebound stratified by quartiles of logarithm of the parameters $\beta$ and $\pi$ (with standard viral dynamics model). Each boxplot shows the median, 25% and 75% quartile. Outliers denoted + are further than 1.5 interquartile range from the nearest end of the box (*boxplot*, MATLAB 2021b). (A) Treatment initiated two days after infection. (B) Treatment initiated three days after infection. (C) The approximated range of each quartile for $\log_{10} \beta$ and $\log_{10} \pi$.

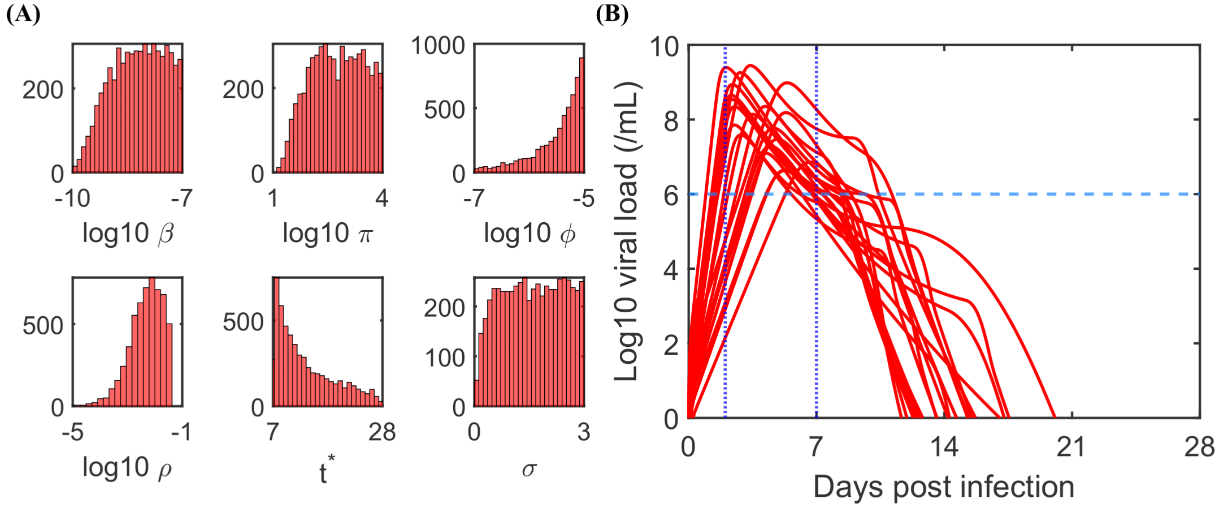


**Fig. S8.** (A) Distribution of accepted parameters – separated by whether the patients rebound or not in the absence of Paxlovid - for the IR model. (B) 20 samples of accepted trajectories in each scenario. The vertical blue dotted lines and the horizontal blue dashed line are the constraints on the viral load peak.

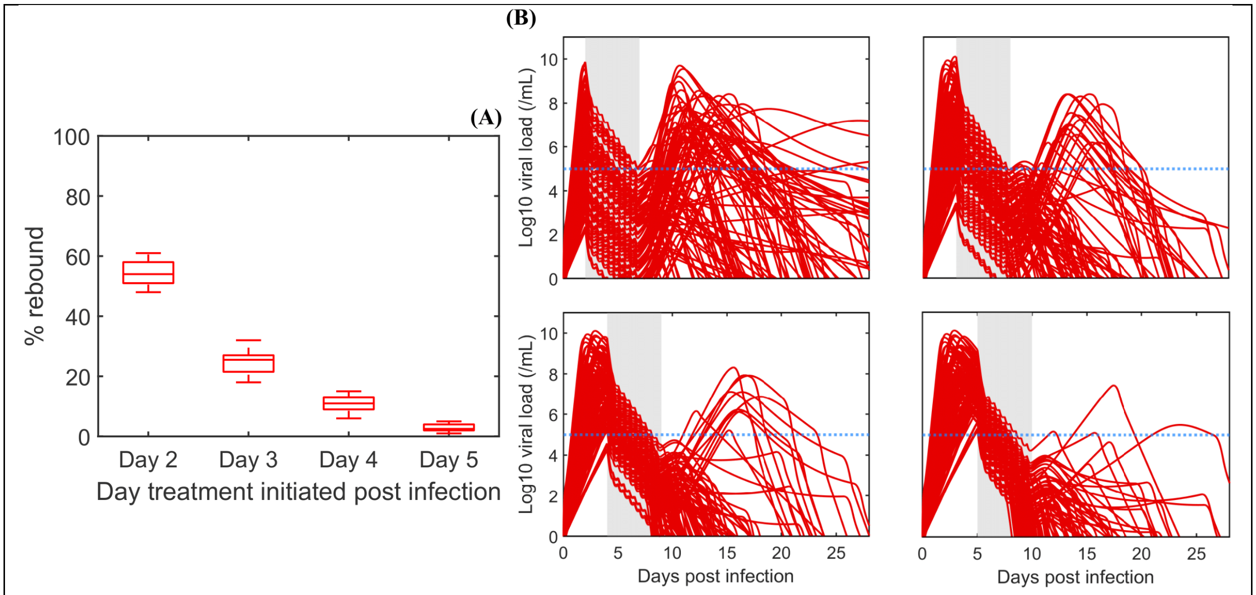


**Fig. S9.** Percentage of rebounds relative to the timing of treatment –immune response model. (A) Percentage of rebounds from 20 runs with 100 samples for each hypothetical treatment time. (B) Examples of rebound from one run with 100 samples. The same 100 samples are used for these examples. The horizontal blue dashed line is the viral load threshold used to define rebound.

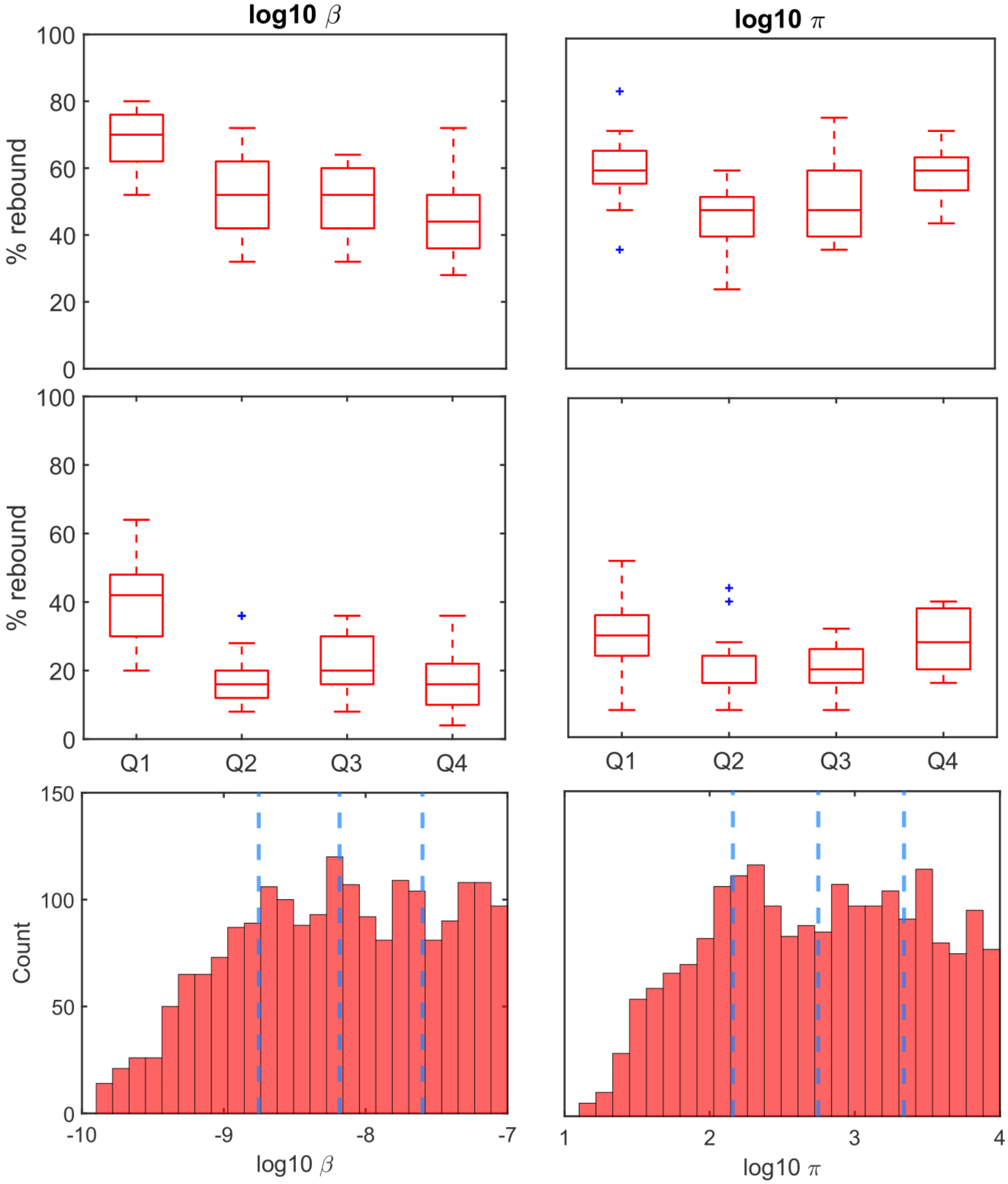


**Fig. S10.** Percentage of predicted *in silico* patients with rebound stratified by quartiles of logarithm of the parameters $\beta$ and $\pi$ (with IR model). Each boxplot shows the median, 25% and 75% quartile. Outliers denoted + are further than 1.5 times the interquartile range from the nearest end of the box (*boxplot*, MATLAB 2021b). (A) Treatment initiated two days after infection. (B) Treatment initiated three days after infection. (C) The approximate range of each quartile for $\log_{10} \beta$ and $\log_{10} \pi$.

| | SSE | | AIC | |
|---|---|---|---|---|
| | Standard | IR | Standard | IR |
| **PATIENT 1** | 3.70 | **2.78** | **-4.00** | -2.58 |
| **PATIENT 2** | **9.70e-7** | 1.34 | **-36.83** | 9.58 |
| **PATIENT 3** | 2.84 | **0.26** | 5.18 | **-2.73** |

**Table S1.** Model fit comparison. Sum of squared error (SSE) and the Akaike Information Criterion score (AIC) for the standard and IR models.

| | *Range* | *Unit* |
|---|---|---|
| $\boldsymbol{\beta}$ | $[10^{-10}, 10^{-7}]$ | mL RNA $\text{copy}^{-1}\text{day}^{-1}$ |
| $\boldsymbol{\pi}$ | $[10^{1}, 10^{4}]$ | copies $\text{mL}^{-1}\text{day}^{-1}$ |
| $\boldsymbol{t}^{*}$ | $[7, 28]$ | days |
| $\boldsymbol{\sigma}$ | $[0, 3]$ | $\text{day}^{-1}$ |
| $\boldsymbol{\phi}$ | $[10^{-7}, 10^{-5}]$ | $\text{cells}^{-1}\text{day}^{-1}$ |
| $\boldsymbol{\rho}$ | $[0, 0.04]$ | $\text{day}^{-1}$ |

**Table S2.** Parameter ranges for the in silico simulation experiments.

Patient 1:

| DPS | 1 | 3 | 5 | 7 | 9 | 11 | 12 | 14 | 18 |
|---|---|---|---|---|---|---|---|---|---|
| Log VL | 8.03 | 6.49 | 5.51 | 5.87 | 9.54 | 8.38 | 7.05 | 6.28 | 4.92 |

The conversion from Ct counts given in ref (8) to viral loads for patients 2 and 3 were kindly provided by David Ho as:

Patient 2:

| DPS | 2 | 10 | 12 |
|---|---|---|---|
| Log VL | 6.27 | 6.41 | 3.38 |

Patient 3:

| DPS | 1 | 4 | 9 | 11 | 13 |
|---|---|---|---|---|---|
| Log VL | 8.19 | 5.41 | 7.42 | 7.18 | 3.87 |

DPS: days post symptom onset. VL: log10 viral RNA copies/mL

**Data S1.** Patient VL data.
